# Comparison of Nanopore with Illumina Whole Genome Assemblies of the Epstein-Barr Virus in Burkitt Lymphoma

**DOI:** 10.1101/2025.02.21.25322471

**Authors:** Isaac E. Kim, Abebe A. Fola, Enrique Puig, Titus K. Maina, Sin Ting Hui, Hongyu Ma, Kaleb Zuckerman, Eddy Agwati, Alec Leonetti, Rebecca Crudale, Micah A. Luftig, Ann M. Moormann, Cliff Oduor, Jeffrey A. Bailey

**Author notes:** **Address all correspondence to:** Jeffrey A. Bailey, MD, PhD, Mencoff Family Associate Professor of Translational Research, Associate Professor of Pathology and Laboratory Medicine, Box G-E5, Providence, RI 02912, USA. **Author Contact Information:** Isaac E. Kim, Jr., Abebe A. Fola, Enrique Puig, Titus K. Maina, Sin Ting Hui, Hongyu Ma, Kaleb Zuckerman, Eddy O. Agwati, Alec Leonetti, Rebecca Crudale, Micah A. Luftig n M. Moormann, Cliff Oduor ffrey A. Bailey. **Grant Funding:** This study was funded by two grants from the National Institutes of Health (R01 CA189806 and R01 CA234348) and the Brown University Blavatnik Family Graduate Fellowship in Biology and Medicine.

## Abstract

Endemic Burkitt lymphoma (eBL) is one of the most prevalent cancer in children in sub-Saharan Africa, and while prior studies have found that Epstein-Barr virus (EBV) type and variation may alter the tumor driver genes necessary for tumor survival, the precise relationship between EBV variation and EBV-associated tumorigenesis remains unclear due to lack of scalable, cost-effective, viral whole-genome sequencing from tumor samples. This study introduces a rapid and cost-effective method of enriching, sequencing, and assembling accurate EBV genomes in BL tumor cell lines through a combination of selective whole genome amplification (sWGA) and subsequent 2-tube multiplex polymerase chain reaction along with long-read sequencing with a portable sequencer. The method was optimized across a range of parameters to yield a high percentage of EBV reads and sufficient coverage across the EBV genome except for large repeat regions. After optimization, we applied our method to sequence 18 cell lines and 3 patient tumors from fine needle biopsies and assembled them with median coverages of 99.62 and 99.68%, respectively. The assemblies showed high concordance (99.61% similarity) to available Illumina-based assemblies. The improved method and assembly pipeline will allow for better understanding of EBV variation in relation to BL and is applicable more broadly for translational research studies, especially useful for laboratories in Africa where eBL is most widespread.

## INTRODUCTION

In 1958, Dr. Denis Burkitt observed a consistent pattern of jaw tumors in Ugandan children, leading to the discovery of Burkitt lymphoma (BL).^1^ Just six years later, Drs. Anthony Epstein and Yvonne Barr discovered the Epstein-Barr virus (EBV) in BL tumor cells, making EBV the first human oncovirus and BL the first viral-associated cancer.^2^ BL has traditionally been categorized into three clinical subtypes primarily based on geography: endemic (eBL) generally found in Africa, sporadic (sBL) found in North America and Europe, and immunocompromised-associated.^3^ Molecular profiling suggests significant difference based on EBV status with EBV-positive BL tumors having higher gene expression levels of activation-induced cytidine deaminase (*AICDA*).^4–8^ Overall, this suggests that BL may be better molecularly defined by EBV’s presence rather than geographic or immunosuppression status of the patient.

EBV also harbors extensive variation, which may affect the tumor’s ability to evade the immune system and hinder apoptosis.^9,10^ While certain EBV variants have been found to increase risk for cancers including nasopharyngeal carcinoma and the nasal type of extranodal NK/T-cell lymphoma,^11^ the exact relationship between EBV variation and tumorigenesis in BL remains more uncertain. Additionally, EBV is classified into type 1 and type 2 based on extensive divergence between *EBNA2* and *EBNA3* genes, which code for essential latent antigens in tumorigenesis, and different studies have shown that EBV type and variation may alter the driver genes necessary for tumor survival.^12–14^ Prior studies discovered six potential variants across four viral genes with EBV type stratification that were enriched in eBL patients and biologically significant.^14^ For example, the two *EBNA1* variants are located on the chromosome-binding domains of the EBNA1 protein, which helps the virus episome bind to the host’s chromosome.^15,16^

Despite EBV’s central role in BL and other diseases, interrogating the viral genome has numerous challenges. First, EBV’s genome (171,823 bp) is approximately 20,000 times smaller than that of humans, and tumors may contain as low as 1 EBV copy/cell.^17^ This makes interrogating the virus limited to targeted sequencing a small proportion of the genome or dependent upon whole genome enrichment methods such as hybrid capture. These enrichment methods require additional expensive library preparation reagents such as RNA baits and significant time.^14^ Second, the virus genome is GC-rich (59.5%^18^), and there are read coverage biases related to GC content in most sequencing and enrichment methods, meaning that the virus is often lost with standard whole genome amplification methods.^19,20^ Third, Illumina sequencing reads are limited in length to 2x300 base pairs (bps), which makes it challenging to traverse across the repetitive regions of the EBV genome. As a result, current viral DNA enrichment methods avoid subsequent analysis of repeat regions larger than 100 nucleotides.^14^ Fourth, the laboratory infrastructure and costs associated with Illumina sequencing machines diminish its accessibility for laboratories in resource-limited areas such as those in Africa where BL is most prevalent. Selective whole genome amplification (sWGA), which seeks to amplify the genome of a target organism amidst the background of another organism, has previously been explored in terms of enriching EBV in BL samples.^14,21^

However, current existing sWGA methods for EBV use the old Phi29™ DNA polymerase, which takes up to 16 hours and only works in low temperature, resulting in lower amplification specificity^14^ and suggesting that better, robust, and more selective target amplification methods are needed to selectively amplify and sequence the EBV genome.

Recently, the advent of Oxford Nanopore Technologies (ONT) has provided a promising avenue for improved sequencing of EBV. Over the last decade, ONT has made significant advances in their nanopore single-molecule sequencers, which have advantages of portability and longer-read lengths. Sample library preparation and the sequencing itself are relatively simple, allowing a general molecular biologist to perform sequencing from beginning to end. The output of the sequencing has also improved in terms of both quality and quantity.^22^ Furthermore, ONT has real-time sequencing capacity, meaning that sequencing can be adjusted and stopped at any point. This ability enables researchers to easily scale from sequencing. ONT sequencing has already been leveraged for genome sequencing of other viruses including SARS-CoV-2.^23,24^ In the future, ONT could help physicians quickly diagnose and treat patients in resource-limited areas.^25^ Altogether, the portability, cost, and real-time sequencing of ONT make it especially attractive for laboratories in developing countries.^26,27^

We have previously developed sensitive enrichment, but it was relatively costly due to secondary hybrid capture.^14^ Towards being able to sequence EBV from tumors in the countries where eBL is endemic, we sought to improve the efficiency of sWGA in combination with ONT sequencing. Additionally, the long-established Phi29™ DNA polymerase enzyme used in current sWGA reactions takes up to 16 hours for incubation and only works in low temperature, resulting in lower amplification specificity^14^. Recently, Thermo Fisher Scientific developed the EquiPhi29™ polymerase, which boasts a shorter reaction time of 3 hours, lower GC bias, and higher reaction temperature, potentially leading to better amplification specificity for EBV.^20^ Therefore, this study aimed to couple sWGA and long-read sequencing to develop a rapid and cost-effective method for targeted whole genome sequencing of EBV from tumor samples that would be easily deployable in Africa.

## MATERIALS AND METHODS

### Tumor Cell Lines

Nine standard BL cell lines were selectively amplified and sequenced including Akata, Akata-GFP (Takada), Daudi, Jijoye, Makau, Mutu I, Namalwa, Raji, and Wewak2. Of these, Akata and Akata-GFP (Takada) were provided by Kenzo Takada (Hokkaido University) ^28,29^, Daudi, Jijoye,

Raji, and Namalwa from the American Type Culture Collection (ATCC) ^30–32^, Mutu I from Jeff Cohen (National Cancer Institute) ^33^, and Makau and Wewak2 from Paul Farrell (Imperial College London). Per established protocols, the cells were preserved at 37°C in a humidified 5% CO_2_ incubator in complete RPMI-1640 medium (Gibco) along with antibiotics (100 U/mL penicillin and 100 μg/mL streptomycin) and 10% heat-inactivated FBS (Fetal Bovine Serum) (Gibco).

The remaining nine BL cell lines (BL717, BL719, BL720, BL725, BL740, BL760, MBL118, MBL120, and MBL121) were more recently established from pediatric BL patients in Kenya, while the three tumor samples (MBL144, MBL213, and BL694) were directly acquired from pediatric BL patients. Upon patient enrollment, all the patient-derived tumor samples were collected before induction of chemotherapy, and diagnosis was confirmed by two independent pathologists via flow-cytometry and cyto-pathology.

### Ethical Approval and Sample Collection

Ethical approval for patient-derived tumor samples was obtained from the Scientific and Ethics Review Unit at the Kenya Medical Research Institute, the Institutional Research and Ethics Committee at Moi Teaching and Referral Hospital, Eldoret Kenya and the Institutional Review Board at the University of Massachusetts Chan Medical School, Worcester, USA. Written informed consent was obtained from the parents of the eBL study participants in accordance with the Declaration of Helsinki.

### EBV Quantification and Typing

Quantification of EBV copies/cell for each cell line was conducted via highly sensitive droplet digital PCR (ddPCR) with viral *BALF* and human β-actin gene.^34,35^ DNA from the Namalwa cell line was the EBV-positive control, while DNA from the BL41 was the EBV-negative control. The duplex ddPCR reactions were prepared in a total volume of 20μL, which contained 10μL of ddPCR Supermix for probes (No UTP) (Bio-Rad Laboratories), and 2 sets of each primer and probe combination (0.9μM of primers and 0.25μM of probes). The Bio-Rad Automated Droplet Generator (AutoDG; Bio-Rad Laboratories) was used to ensure consistent droplet generation. After the ddPCR reaction, the number of positive and negative droplets were counted by the QX200TM Droplet reader (Bio-Rad) and EBV viral loads quantified as EBV copies/ng human DNA.

EBV type for the cell lines was determined by mapping the ONT sequencing reads onto the reference genomes of each EBV type, NC_007605 (Type 1) and NC_009334 (Type 2), and then examining which type recruited the most reads at their respective *EBNA2* and *EBNA3* genes.

### Enrichment of EBV

DNA from each cell line was isolated using the Monarch Genomics DNA Purification Kit (NEB). The key principle behind sWGA is finding specific sequences for short primers more common in the target genome (in our case EBV) than other background genomes (i.e. tumor DNA/human genome).^21,36^ Primers are then used in isothermal amplification, allowing faster amplification due to the higher density of priming sites in the target genome. To start our long-read sWGA pipeline, sWGA was performed using Thermo Fisher Scientific’s EquiPhi29 DNA polymerase with a set of EBV-specific 3’-protected oligonucleotides designed per Leichty and Brisson’s protocol.^14,21^ The sWGA reaction was incubated for 3 hours at 45°C followed by inactivation for 10 minutes at 65°C **(Figure 1)**. To cleave DNA branches that would clog the pores on the ONT flow cell and significantly decrease sequencing throughput, up to 1500 ng of the amplified DNA from the sWGA reaction volume was debranched with T7 endonuclease I for 15 minutes at 37°C and purified with Agencourt AMPure XP beads suspended in a custom buffer using a slightly modified ONT protocol. 600 ng of the debranched product was split into six PCR reactions, two for the multiplexed pools of primers tiling across the whole EBV genome per

**Figure 1.**
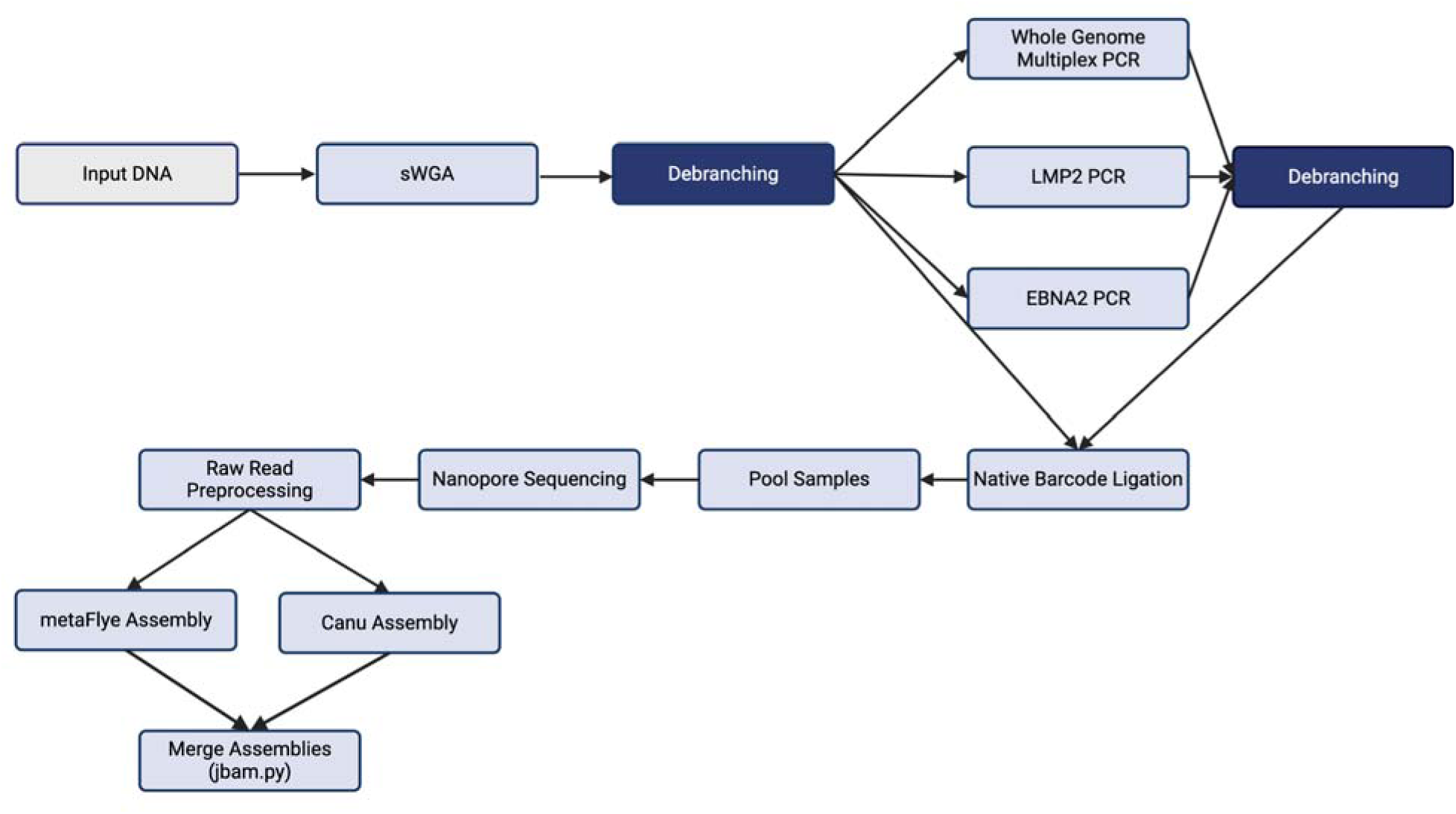
EBV long-read sWGA Pipeline. An initial 12.5 ng of input DNA is amplified with sWGA and then debranching. 600 ng of the debranched product is distributed for whole genome multiplex PCR (200 ng), *LMP2* singleplex PCR (300 ng), and *EBNA2* singleplex PCR (100ng), which is followed by a second round of debranching. The final post-PCR products are consolidated for ONT sequencing, raw read preprocessing, and genome assembly by Canu and metaFlye. The two assemblies are then merged into a single assembly.

Kwok et al. and EBV type 2-specific primers,^14,37^ one for a singleplex EBNA2 primer from Kwok et al., and three for custom primers designed for LMP2 and the ends of the viral genome **(Figure 2).** The Integrated DNA Technology (IDT) and National Center for Biotechnology Information (NCBI) Primer-BLAST software were used to develop these three primer sets. Each PCR reaction required 100 ng of input DNA and KAPA HiFi HotStart ReadyMix and entailed initial denaturation at 95°C for 3 minutes followed by 25 cycles of denaturation at 98°C for 20 seconds, gradient annealing from 58 to 49°C for 15 seconds per degree, and extension at 72°C for 7 minutes, concluded with a final extension at 72°C for 10 minutes. Identical conditions for the six PCR reactions allowed them to be performed on the same thermocycler. These reaction volumes were then debranched for a second time using the same protocol mentioned above to cleave any residual branches that may be present.

**Figure 2.**
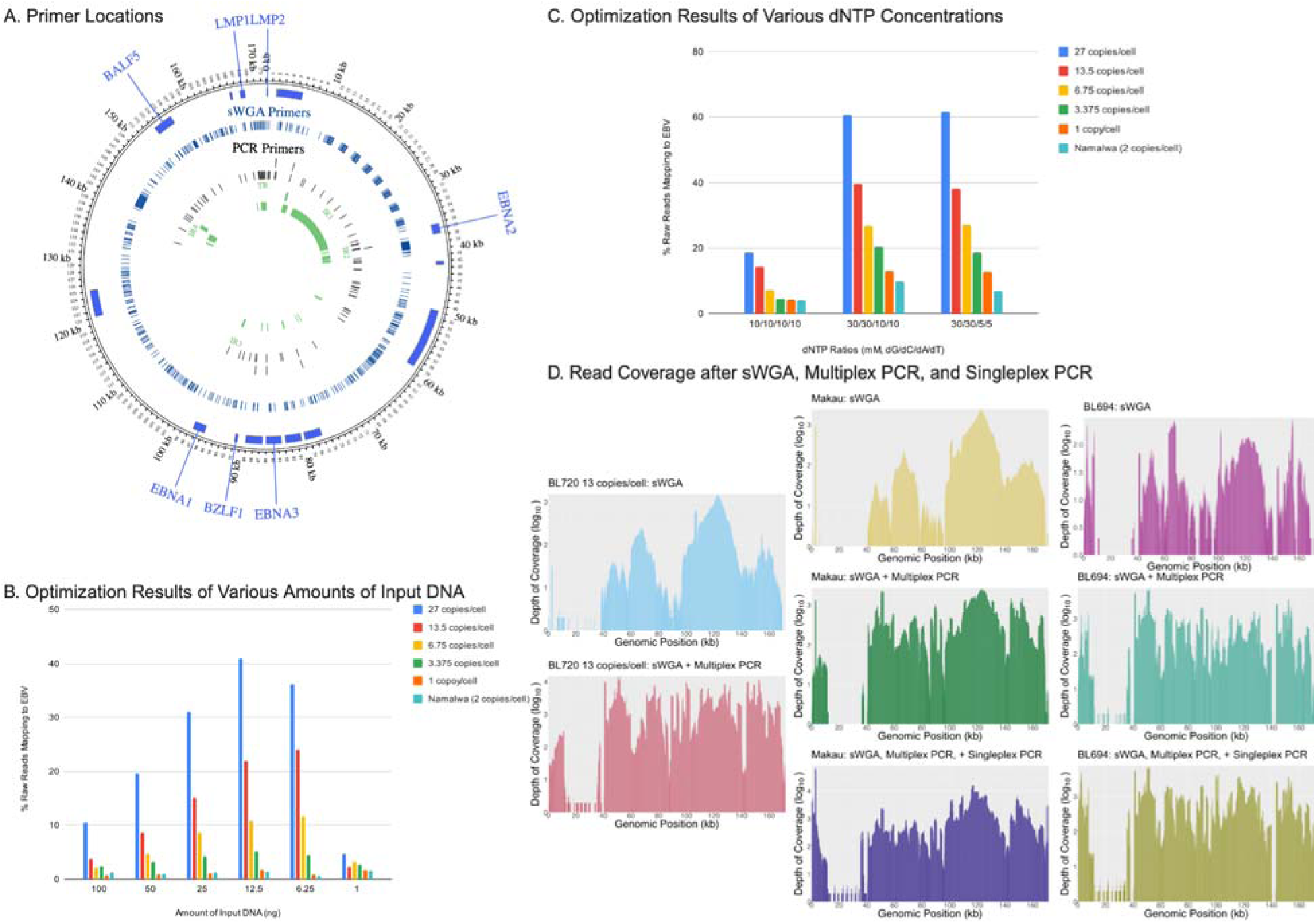
Experimental sWGA and PCR Optimizations. (A) Location of sWGA and PCR Primers on EBV Genome (B) sWGA Optimization results of various amounts of input DNA measured by the percentage of total reads mapping to EBV (C) sWGA Optimization results of various dNTP concentrations measured by the percentage of total reads mapping to EBV (D) Read coverage after sWGA, sWGA/multiplex PCR, and sWGA/multiplex PCR/singleplex PCR for BL720 dilution 13 copies/cell, cell line Makau, and patient tumor sample BL694.

### ONT Sequencing Library Preparation

For ONT sequencing library preparation, 400 ng of each debranched product (post-sWGA, combined post-multiplex PCR pools 1 and 2, and post-EBNA2 and LMP2 singleplex PCR) were prepared, barcoded, and sequenced separately. ONT sequencing library preparation was based on the Native Barcoding Kit 24 V14 protocol, which included DNA repair and end-prep, native 24-barcode ligation (SQK-NBD114.24), and adapter ligation and clean-up with the R10.4.1 chemistry kit. The final elution was sequenced on a PromethION flow cell for a targeted whole genome sequencing output of 400x or 137 megabases per sample (400 *172,000 (bps) / (50% EBV)). The full protocol from sWGA to ONT sequencing is available in the supplement (**Supplemental Material 1**).

### Sequence Preprocessing and Genome Assembly

The MinKNOW software then processed the data based on magnitude changes in the ion current flow through the pores.^38^ The electronic raw signals of the reads from the sequencer were translated into bases with ONT’s Guppy 6.4.8 high accuracy base-calling software. Adapter sequences were then removed from the reads with Cutadapt (version 4.7).^39^ If adapter sequences were not found in a given read, 10 bases were trimmed from both ends of the read. Median read depth was calculated with samtools v1.6,^40^ while read depth at repeat regions was calculated with Mosdepth.^41^ Processed sequencing reads were then *de novo* assembled using Canu and metaFlye for the reads and Canu for the *EBNA2*-mapping reads with modified parameters to ensure that all reads were used (Canu for all reads: maxInputCoverage of 1,000,000,000, minReadLength of 200, and minOverLapLength of 100; metaFlye for all reads: --nano-raw and --meta; Canu for *EBNA2*-mapping reads: maxInputCoverage of 1,000,000,000, minReadLength of 40, and minOverLapLength of 40).^42,43^ metaFlye discarded reads shorter than 1000 bp. Since *EBNA2* is only 1463 bps long and wedged in between repeat regions, many EBNA2-matching reads were discarded in metaFlye.^40,41^ Furthermore, most *EBNA2* sequencing reads are contained within a larger, encompassing read. As a result, per recommendation of the Canu authors, the *EBNA2* region was specifically and exclusively assembled by taking the longest trimmed read with a further modified Canu (maxInputCoverage of 9,999,999, minReadLength of 40, and minOverLapLength of 40). To ensure both high accuracy and contiguity, two long-read assembly algorithms were selected: Canu and metaFlye. Roughly, Canu initially corrects and trims all reads and then assembles them, leading to more accurate but possibly shorter assemblies.^43^ In contrast, metaFlye first assembles the reads via a repeat graph and then corrects the assembly, potentially resulting in longer assemblies but slightly lower accuracy.^42,44^ The resulting contigs were then polished and merged using a custom, reference-based script that determined the final assembly based on the mapped bam files containing the original reads and contigs. Note that this script sought to elucidate the major viral haplotype, as multiple viral strains appear to be very rare in tumors.^45^ By leveraging both algorithms and then polishing and merging the assemblies with an in-house script, high accuracy and contiguity were achieved. Mappings and sorting of the bam file were performed with minimap2 and samtools, respectively.^46,47^ The contigs were scaffolded based on the reference genomes NC_007605 and NC_009334 using RagTag.^48^ The assemblies were masked at repeat regions using RepeatMasker (version 4.1.6).^49^ Finally, assemblies were annotated for features with VAPiD version 1.2.^50^

### Illumina Assembly Comparison

From the 18 cell lines sequenced with ONT, 14 publicly available, Illumina-based assemblies of the cell lines were obtained.^14^ Using the Quality Assessment Tool for Genome Assemblies (QUAST),^51^ ONT assemblies were then compared to the Illumina assemblies to measure similarity with the longer genome serving as the reference. Enrichment for EBV was calculated with the following equation accounting for the lengths of the EBV reference (171823 bp) and human genome (3298912062 bp haploid) as well as the number of EBV copies/cell in each sample: [ [(# EBV nucleotides) / (# EBV nucleotides + # Human nucleotides)] / [(# EBV copies/cell * 171823) / (# EBV copies/cell * 171823 + 6597824124)]]. Enrichment for human was calculated with the following: [ [(# Human nucleotides) / (# EBV nucleotides + # Human nucleotides)] / [(# EBV copies/cell * 171823) / (# EBV copies/cell * 171823 + 6597824124)]]. To roughly calculate the shortest length of the contig (N50) and lowest number of contigs (L50) needed to assemble at least half of the genome size, ONT assemblies were also mapped to the reference genome NC_007605 and analyzed with QUAST. Assembly coverage at repeat regions was calculated with bedtools v2.31.0.^52^ To determine the significance of a reaction component such as SWGA, multiplex PCR, or singleplex PCR, statistical analyses using Stata/SE 18.0 included the Pearson’s correlation and paired Student’s T-test on the percentage of bases mapping to EBV before and after the reaction component for each given sample and were conducted.^53^

### EBV Phylogenetic Analysis

After masking for repeats, ONT assemblies of all cell lines along with their Illumina counterparts were aligned across the entire genome with multiple sequence alignment (msa) using MAFFT v7.511 with the nwildcard parameter,^54^ from which a phylogenetic tree was created using the neighbor-joining method with the Jukes-Cantor substitution model bootstrapped 1000 times. The resulting Newick tree files were re-formatted in RStudio with the R packages tidyverse v2.0.0, ggtree v3.8.2, ggtreeExtra v1.10.0, and ggplot2 v3.4.3.^55–59^

## RESULTS

### sWGA was optimized for amount of input DNA, dNTP composition, debranching, and reaction temperature

Initial optimizations were conducted using the BL720 cell line (27 EBV copies/cell) and subsequent mixtures after dilution to create viral dilutions with EBV-negative human DNA representing 27, 13.5, 6.75, 3.375, and 1 EBV copies / human cell (**Supplementary Table 1**). In these optimizations, we first sought to optimize the sWGA reaction. To maximize the relative yield of EBV to human DNA, various aspects of the selective amplification pipeline were tested using BL720 cell line DNA (Table 1). Stock sample as well as all serial dilutions with human control DNA (13.5, 6.75, 3.375, and 1 copy/cell) were confirmed by EBV qPCR (**Supplementary Table 1**). For sWGA, the highest percentage of EBV-mapping reads for all BL720 dilutions was obtained with an input DNA volume of 12.5 ng (**Figure 1**, **Supplementary Table 3**). More than 12.5 ng of input DNA (100, 50, and 25 ng) had a slightly decreased yield, while less than 12.5 (6.25 and 1 ng) did not reach the enzyme’s maximum capacity for amplification. Additionally, since EBV has a high GC content (59.5%),^18^ the relative concentrations of dNTPs were explored. Similar to prior findings,^14^ the percentage of reads mapping to EBV was maximized with greater ratio of dGTP and dCTP. While 30/30/5/5 and 30/30/10/10 of dG/dC/dA/dT achieved similar percentages, 30/30/5/5 was selected, because it achieved a slightly higher percentage at the highest EBV concentration for BL720 at 27 copies/cell, and most patient tumor samples have at least 27 EBV copies/cell.

**Table 1.** Patient Characteristics.

| Category | No. (%) |
| --- | --- |
| **<u>BL Cell Lines</u>** |  |
| **Sex** |  |
| Male | 9 (60) |
| Female | 6 (40) |
| **Median Age (years)** | 6 |
| **Tumor Site** |  |
| Jaw | 7 (50) |
| Neck | 1 (7.14) |
| Orbit | 1 (7.14) |
| Abdomen | 5 (35.71) |
| **Median EBV Copies/Cell** | 19 |
| **EBV Type** |  |
| Type 1 | 14 (77.78) |
| Type 2 | 4 (22.22) |
| **<u>Patient Tumor Fine Needle Aspirates</u>** |  |
| **Sex** |  |
| Male | 1 (33.33) |
| Female | 2 (66.67) |
| **Median Age (years)** | 9 |
| **Tumor Site** |  |
| Abdomen | 2 (100) |
| **Median EBV Copies/Cell** | N/A |
| **EBV Type** |  |
| Type 1 | 3 (100) |
| Type 2 | 0 (0) |

\*Measured by ddPCR

Next, since the EquiPhi29™ polymerase amplifies DNA through multiple displacement amplification, it produces DNA branches that can clog pores on the ONT flow cell.^60^ In terms of sequence optimization, the sWGA product underwent a debranching reaction with the T7 endonuclease to prevent clogging and maximize sequencing output. There are currently two debranching protocols for ONT sequencing: one from New England Biolabs (NEB)^61^ and the other from ONT (**Supplemental Material 1**). The main difference between the two protocols was the amount of input DNA with NEB’s protocol requiring 200 ng and ONT’s requiring 1500 ng. Both debranching reactions yielded a lower amount of DNA compared to the initial input, and subsequent reactions in the pipeline such as PCR required greater than 200 ng of DNA after debranching. As a result, we adopted ONT’s debranching protocol for our pipeline. Initial optimized output of the debranched sWGA from sequencing, hereafter described as post-sWGA, showed a median read length of 1449 bps, significant 1410-fold enrichment of EBV, 0.83-fold enrichment of human, and the total amount of DNA increased from 12.5 ng to 2652 ng (**Table 2**, **Supplementary Table 4**). Ultimately, the sWGA reaction was finalized in a 20 μL reaction with 12.5 ng of total input DNA and 30/30/5/5 dG/dC/dA/dTTP.

**Table 2.**
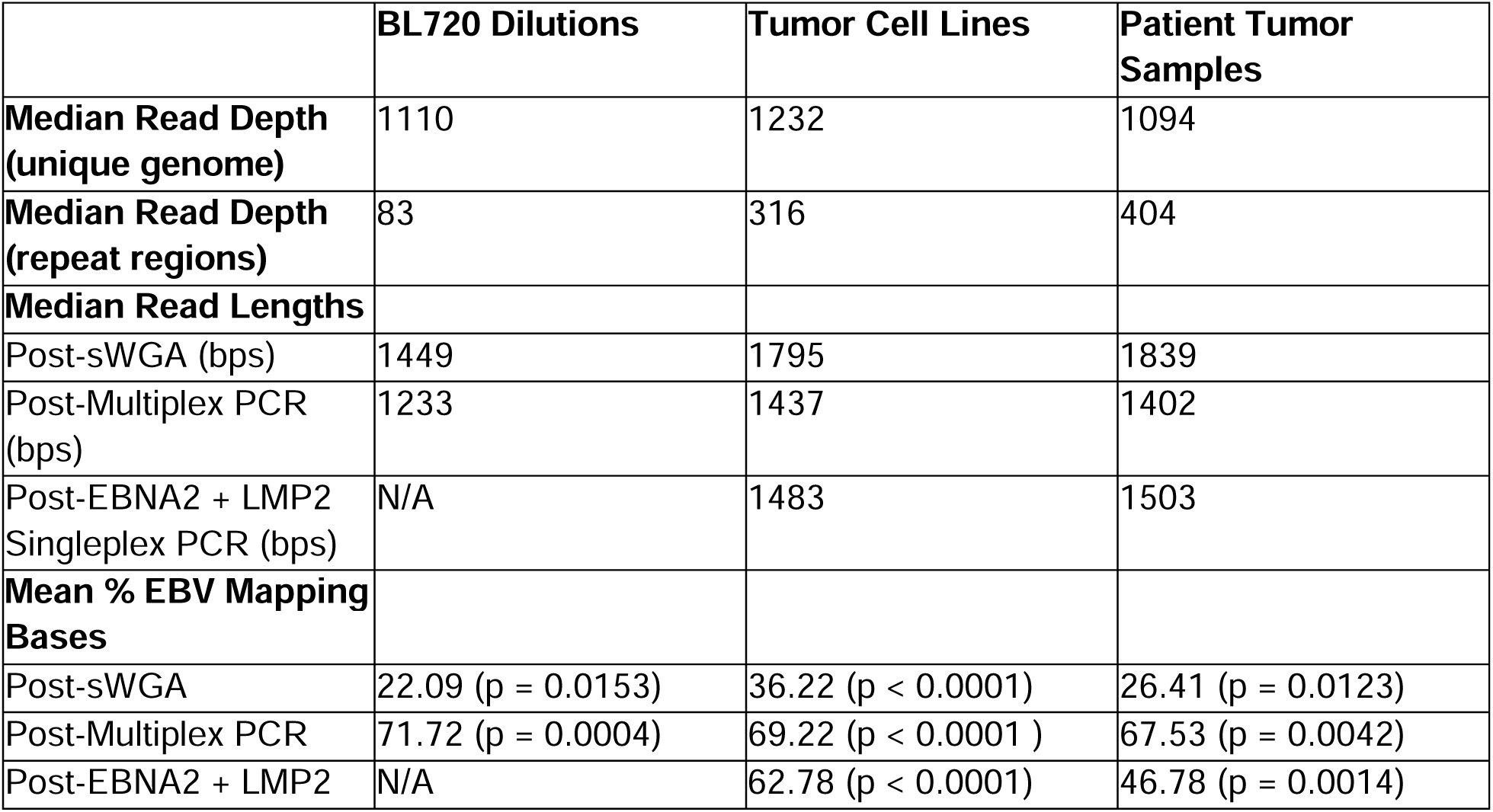
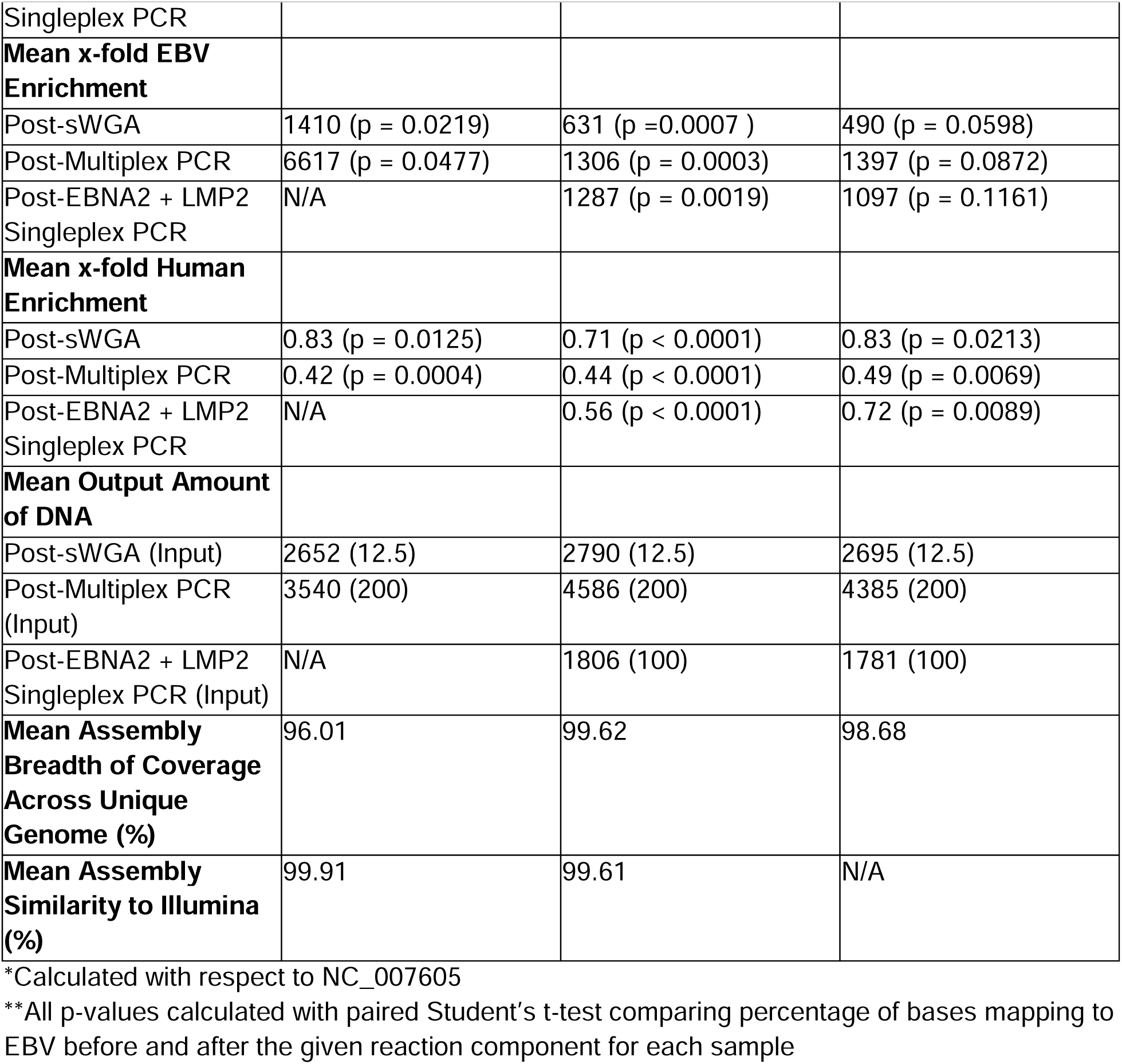
Read and Assembly Statistics.

|  | **BL720 Dilutions** | **Tumor Cell Lines** | **Patient Tumor Samples** |
| --- | --- | --- | --- |
| **Median Read Depth (unique genome)** | 1110 | 1232 | 1094 |
| **Median Read Depth (repeat regions)** | 83 | 316 | 404 |
| **Median Read Lengths** |  |  |  |
| Post-sWGA (bps) | 1449 | 1795 | 1839 |
| Post-Multiplex PCR (bps) | 1233 | 1437 | 1402 |
| Post-EBNA2 + LMP2 Singleplex PCR (bps) | N/A | 1483 | 1503 |
| **Mean % EBV Mapping Bases** |  |  |  |
| Post-sWGA | 22.09 (p = 0.0153) | 36.22 (p < 0.0001) | 26.41 (p = 0.0123) |
| Post-Multiplex PCR | 71.72 (p = 0.0004) | 69.22 (p < 0.0001 ) | 67.53 (p = 0.0042) |
| Post-EBNA2 + LMP2 | N/A | 62.78 (p < 0.0001) | 46.78 (p = 0.0014) |
| Singleplex PCR |  |  |  |
| **Mean x-fold EBV Enrichment** |  |  |  |
| Post-sWGA | 1410 (p = 0.0219) | 631 (p = 0.0007 ) | 490 (p = 0.0598) |
| Post-Multiplex PCR | 6617 (p = 0.0477) | 1306 (p = 0.0003) | 1397 (p = 0.0872) |
| Post-EBNA2 + LMP2 Singleplex PCR | N/A | 1287 (p = 0.0019) | 1097 (p = 0.1161) |
| **Mean x-fold Human Enrichment** |  |  |  |
| Post-sWGA | 0.83 (p = 0.0125) | 0.71 (p < 0.0001) | 0.83 (p = 0.0213) |
| Post-Multiplex PCR | 0.42 (p = 0.0004) | 0.44 (p < 0.0001) | 0.49 (p = 0.0069) |
| Post-EBNA2 + LMP2 Singleplex PCR | N/A | 0.56 (p < 0.0001) | 0.72 (p = 0.0089) |
| **Mean Output Amount of DNA** |  |  |  |
| Post-sWGA (Input) | 2652 (12.5) | 2790 (12.5) | 2695 (12.5) |
| Post-Multiplex PCR (Input) | 3540 (200) | 4586 (200) | 4385 (200) |
| Post-EBNA2 + LMP2 Singleplex PCR (Input) | N/A | 1806 (100) | 1781 (100) |
| **Mean Assembly Breadth of Coverage Across Unique Genome (%)** | 96.01 | 99.62 | 98.68 |
| **Mean Assembly Similarity to Illumina (%)** | 99.91 | 99.61 | N/A |

\*Calculated with respect to NC\_007605
\*\*All p-values calculated with paired Student's t-test comparing percentage of bases mapping to EBV before and after the given reaction component for each sample

### Supplementation with region-specific multiplex PCR to optimize coverage

Optimized sWGA did not show uniform amplification with an overrepresentation of the region spanning 110,000 to 130,000 bases in the reference genome (**Figure 2**). As a result, we added two-tube multiplex PCR leveraging previous primers that targeted all parts of the EBV genome except for bases 110,000-130,000 to the post-sWGA product. These PCR reactions were optimized with 200 ng of input DNA (100 ng for each reaction) and a gradient annealing temperature ranging from 58° to 49°C to accommodate for the multiple primers and their varying melting temperatures. From 200 ng of input DNA after sWGA, the PCR reactions, henceforward referred to as post-multiplex PCR, outputted median read lengths of 1233 bps and an average of 3540 ng of DNA across all BL720 dilutions (**Table 2**). Post-multiplex PCR provided more uniform coverage (Figure 2) with a 6617-fold increase in enrichment of EBV (*p*=0.05) and 0.42-fold enrichment of human (*p*<0.01, **Table 2**). Finally, after PCR, we obtained increased sequencing output by debranching the reaction again with T7 endonuclease. Note that we observed significantly decreased sequencing throughput when only doing a single round of debranching for post-multiplex PCR compared to two rounds total with one round for post-sWGA and another for post-multiplex PCR. Overall, all non-repetitive regions of the EBV genome, henceforth referred to as the unique genome, were covered by sequencing reads with a median read depth of 1124 across all measured ranges of EBV copies/cell for BL720 **(Figure 3).** The 14 repeat regions observed a median read depth of 83 for all BL720 dilutions with all repeat regions smaller than 1238 bases being covered across all dilutions (**Table 2**, **Supplementary Table 6**, **Supplementary Table 7**). In contrast, the largest regions (*IR1* at 23,355 bps, *IR4* at 2517 bps, *TR* at 2137 bps, and *IR2* at 1538 bps) did not have read coverage in at least one BL720 dilution (**Supplementary Table 6**, **Supplementary Table 7**). Altogether, across all measured ranges of EBV copies/cell for BL720, the combination of post-sWGA and post-multiplex PCR followed by ONT long-read sequencing demonstrated significant enrichment of EBV relative to human and relatively uniform read coverage across the unique genome (**Table 2**, **Figure 3**).

**Figure 3.**
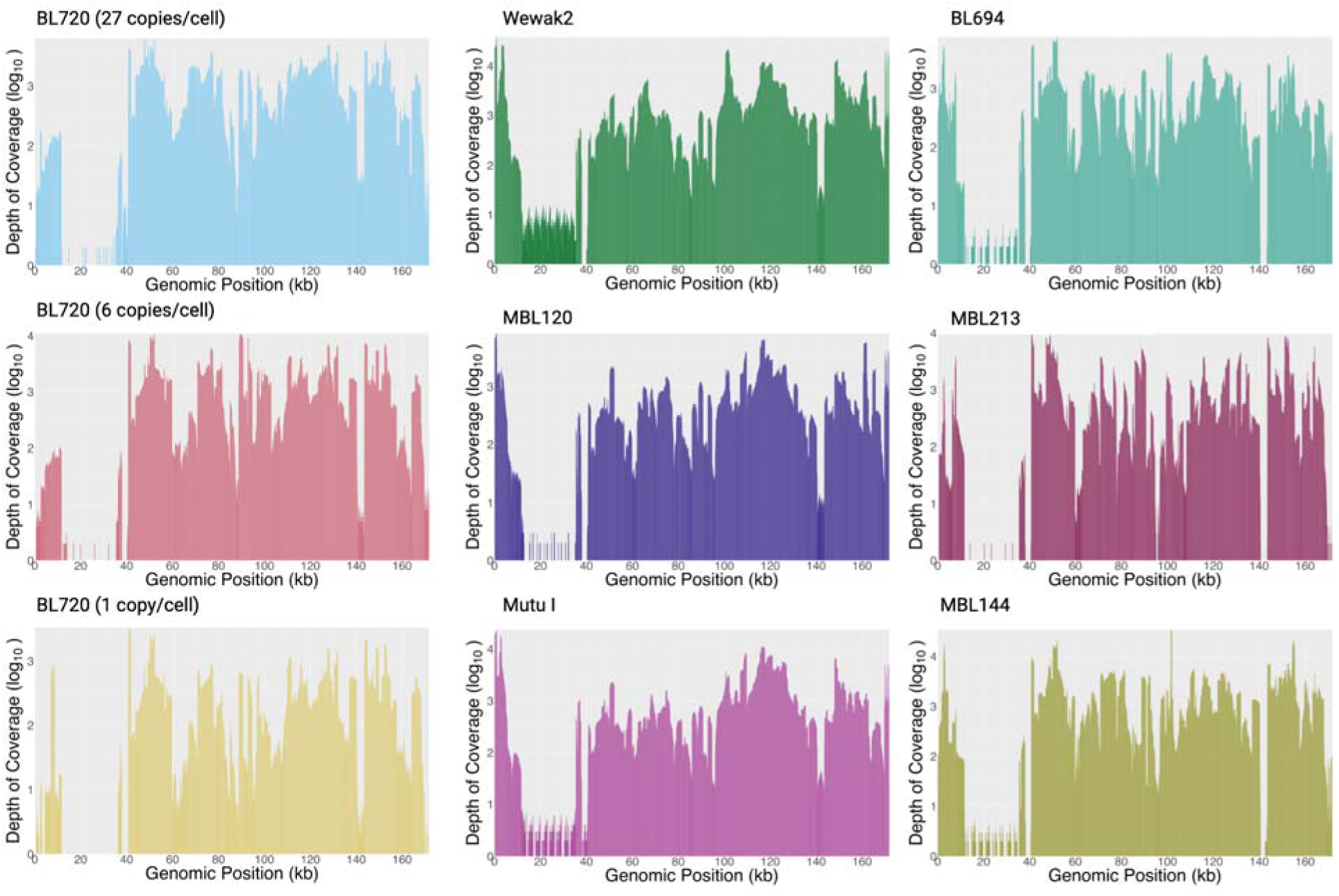
Representative Long Read Coverage. (A) Read Coverage Across Genome of BL720 (27 copies/cell down to 1 copy/cell) (B) Read Coverage Across Genome of Tumor Cell Lines (C) Read Coverage Across Genome of Patient Tumor Samples. Note that EBNA2 is at genomic positions 36216-37679, and the targeted LMP2 region is at 0-1680 and 171772-171823. Created with BioRender.com

### Sequencing of tumor cell lines and further optimization for low coverage in *EBNA2* and LMP2

With our optimized parameters for sWGA, multiplex PCR, and debranching, we sequenced a broader array of BL cell lines (Akata, Akata GFP, Daudi, BL717, BL719, BL725, BL740, BL760, Jijoye, Makau, Mutu I, Namalwa, Raji, Wewak2, MBL118, MBL120, and MBL121). 18 total cell lines were also sequenced, of which 9 were derived from male patients and 6 female (**Table 1**, **Supplementary Table 2**). The original tumor sites included abdomen, jaw, neck, orbit, and ascitic fluid. The median EBV copies/cell for the cell lines was 27 copies/cell (range 1-132) with 77.78% being EBV type 1 and 22.22% type 2. All showed significant fold EBV enrichment (631) and human enrichment (0.71) after sWGA with a DNA yield of 2790 ng and median read length of 1795 bps and 1306-fold EBV enrichment and 0.44-fold human enrichment after multiplex PCR (**Table 2**), yielding 4586 ng of DNA with median read length of 1437 bps. Similar to the BL720 dilutions, all cell lines demonstrated biased coverage at bases 110,000-130,000 after sWGA and more uniform coverage after multiplex PCR (**Figure 2**).

While the BL720 dilutions had sufficient coverage across all genes after sWGA and multiplex PCR, there were regions of consistently lower coverage in the cell lines, specifically in *EBNA2* and *LMP2A/2B*. These regions likely observed relatively lower coverage due to the repeat regions flanking *EBNA2* and the terminal repeat region within *LMP2A/2B*.^62,63^ Since increasing the concentration of PCR primers at these regions in the multiplex PCR reaction did not significantly increase coverage, we designed single primer-pairs (singleplex) PCR amplifying these genes after the initial sWGA, henceforth referred to as post-singleplex PCR. Post-singleplex PCR resulted in 1287-fold EBV enrichment and 0.56-fold human enrichment, a total output of 1806 ng of DNA, median read length of 1483 bps, and increased coverage at *EBNA2* and *LMP2A/2B* compared to the original, unamplified DNA (**Figure 2**, **Figure 3**, **Supplementary Table 4**). For the final coverage, the median read depth was 1232, and there was statistically significant positive correlation between GC content of assemblies and reads mapping to EBV after sWGA (*p*=0.03, Pearson’s correlation; **Supplementary Table 4**). The median read depth for the repeat regions was 316 (**Table 2**). 12 of the 14 repeat regions had read coverage across all cell lines. The two regions that failed included *IR1*, which measures 23,355 bps, and *IR2* at 1538 bps (**Supplementary Table 6**, **Supplementary Table 7**). Note that multiplex PCR and singleplex PCR were performed concurrently, meaning that both reactions used the post-sWGA product (**Figure 1**). Thus, each sample was assigned three barcodes during the native barcode ligation step of ONT’s Native Barcoding Kit 24 V14 protocol. After the native barcode ligation step, however, all barcodes were combined into a single library for adapter ligation and priming and loading the flow cell.

We then applied our final optimizations on the cell lines for three tumor samples, which consisted of one male and two female patients with the two known tumor sites being the abdomen (**Supplementary Table 2**). All contained type 1 EBV. The three patient tumor samples had 26.4%, 67.5%, and 46.8% of total bases mapping to EBV for post-sWGA, post-multiplex PCR, and post-singleplex PCR, respectively (**Table 2**, **Figure 3**). For these patient tumors, the median read depth was 1094, and patterns in read coverage across the genome for post-sWGA, post-multiplex PCR, and post-singleplex PCR resembled those of the cell lines and BL720 dilutions (**Figure 2**, **Figure 3**). Ten of the 14 repeat regions demonstrated read coverage across all patient tumor samples. The four repeat regions that failed were some of the longest repeat regions including *IR1* (23,355 bps), *IR2* (1,538 bps)*, IR4* (2,517 bps), and *TR* (2,137 bps)

### ONT assemblies covered all key genes with L50 of 1 and high identity to Illumina

Across all assemblies, there were no consistent gaps in any genes across the EBV genome (**Figure 4**). For the BL720 sample that was diluted with additional human DNA to generate 1 EBV copy/cell, the EBV assembly covered 93.90% of the unique genome at 99.91% similarity to the Illumina assembly (**Table 2**, **Supplementary Table 8**). In comparison, the undiluted BL720 at 27 copies/cell was sequenced and assembled at 98.60% coverage breadth and 99.86% similarity to the Illumina assembly. Of the 18 cell lines, 14 had Illumina assemblies, which served as the reference genomes for coverage breadth and similarity measurements. All cell line assemblies observed at least 98.60% coverage breadth (**Supplementary Table 8**). The average similarity of the ONT assemblies to the Illumina assemblies was 99.61%. Due to the absence of any prior assemblies, assembly breadth of coverage for the patient tumor samples was estimated by comparing to the EBV type 1 reference genome NC_007605. The estimated assembly breadths of coverage for MBL144, MBL213, and BL694 were 98.72%, 98.71%, and 98.60%, respectively (**Supplementary Table 4**). All BL720 dilution, cell line, and patient sample final assemblies had L50s of 1 with average N50s of 166,059, 170,257, and 168,673, respectively. In 13 of the 21 repeat regions defined in NC_007605 including all repeat regions less than 993 bps except the 183-bp *R* gene, at least 80% of the BL720 dilution assemblies completely covered the region, hereon termed continuous assembly coverage (**Table 3, Supplementary Table 9, Supplementary Table 10)**. Similarly, at least 80% of the cell line assemblies had continuous assembly coverage in 14 repeat regions including all repeats less than 993 bps (**Table 3, Supplementary Table 9, Supplementary Table 10)**. Finally, all patient tumor assemblies had continuous assembly coverage in 12 repeat regions including all regions smaller than 708 bps (**Table 3, Supplementary Table 9, Supplementary Table 10)**.

**Figure 4.**
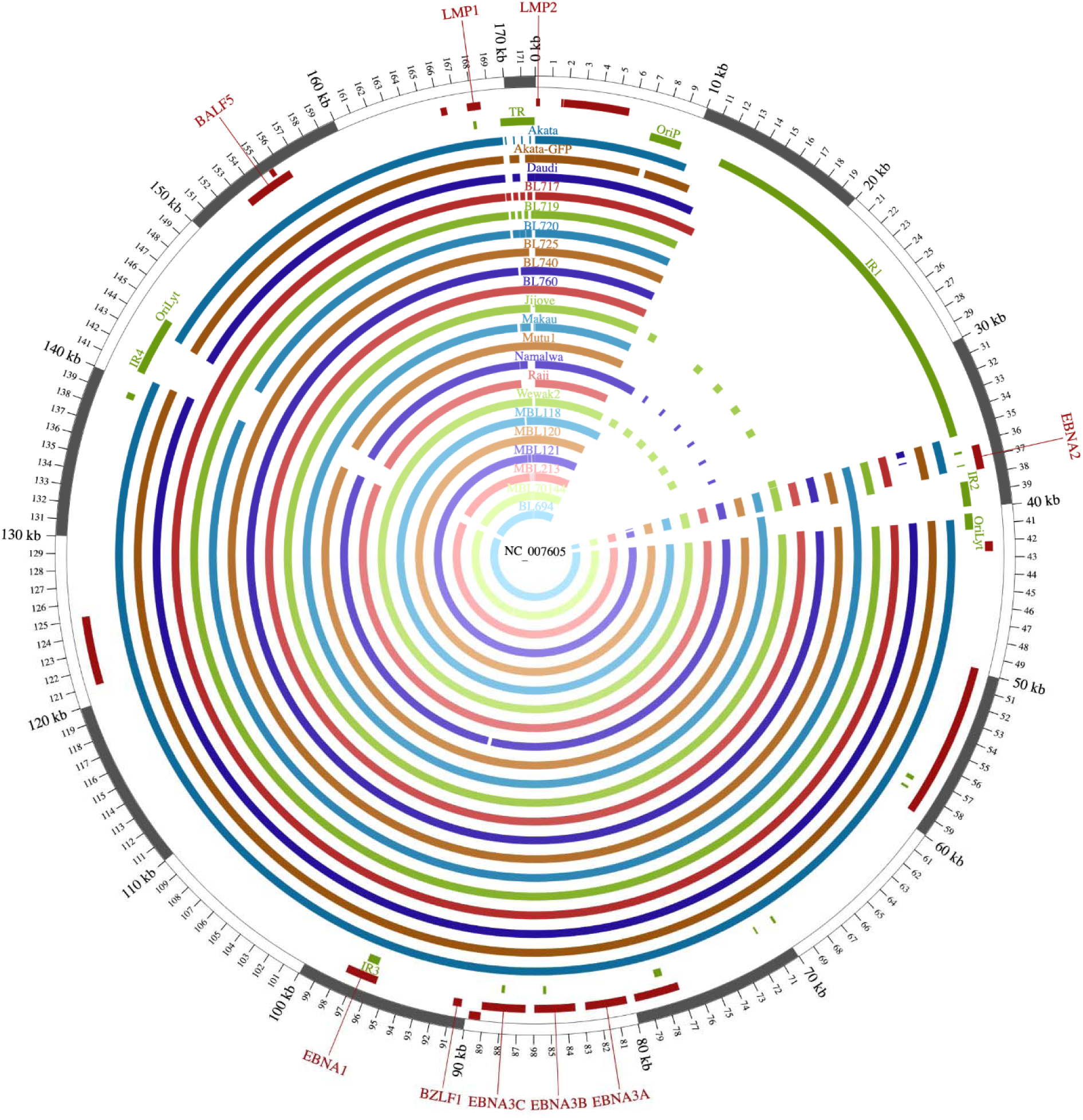
Assemblies Coverage. Assemblies for the 18 tumor cell lines (Akata, Akata GFP, Daudi, BL717, BL719, BL720, BL725, BL740, BL760, Jijoye, Makau, Mutu I, Namalwa, Raji, Wewak2, MBL118, MBL120, and MBL121) and 3 FNA tumor samples (MBL213, MBL144, and BL694) were mapped to the reference genome NC_007605. All assemblies covered all genes (ie *EBNA2, EBNA3, BZLF1, EBNA1, BALF5, LMP1,* and *LMP2*), and the median continuous assembly coverage across all assemblies for repeat regions up to 1000 bps was 100.00%.

**Table 3.**
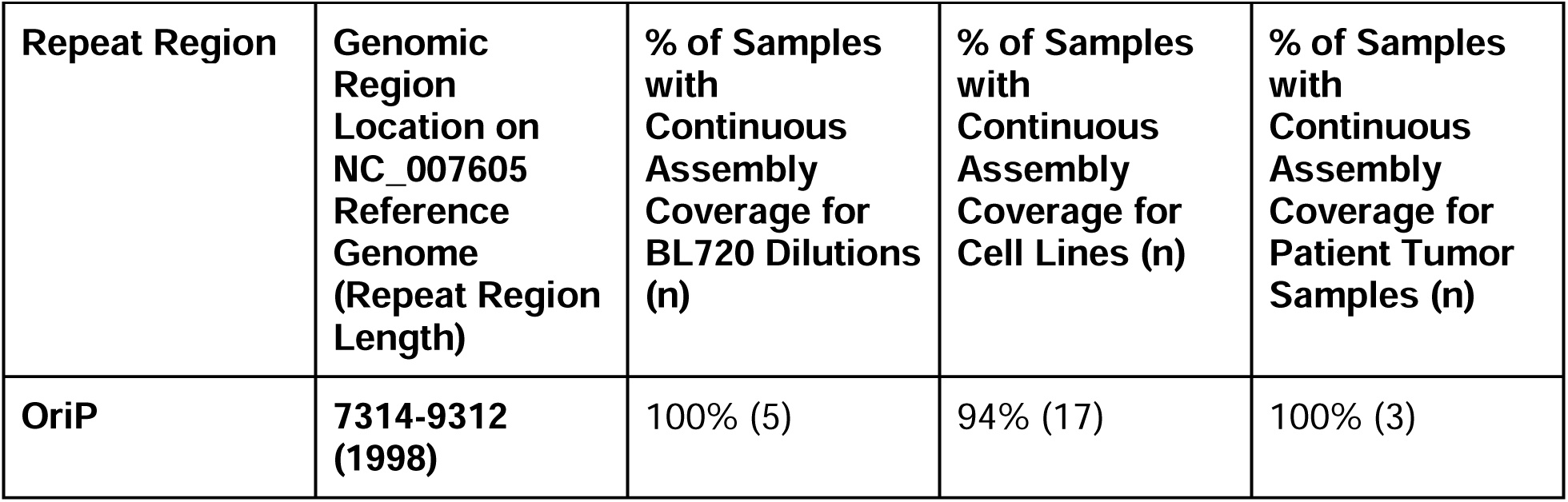
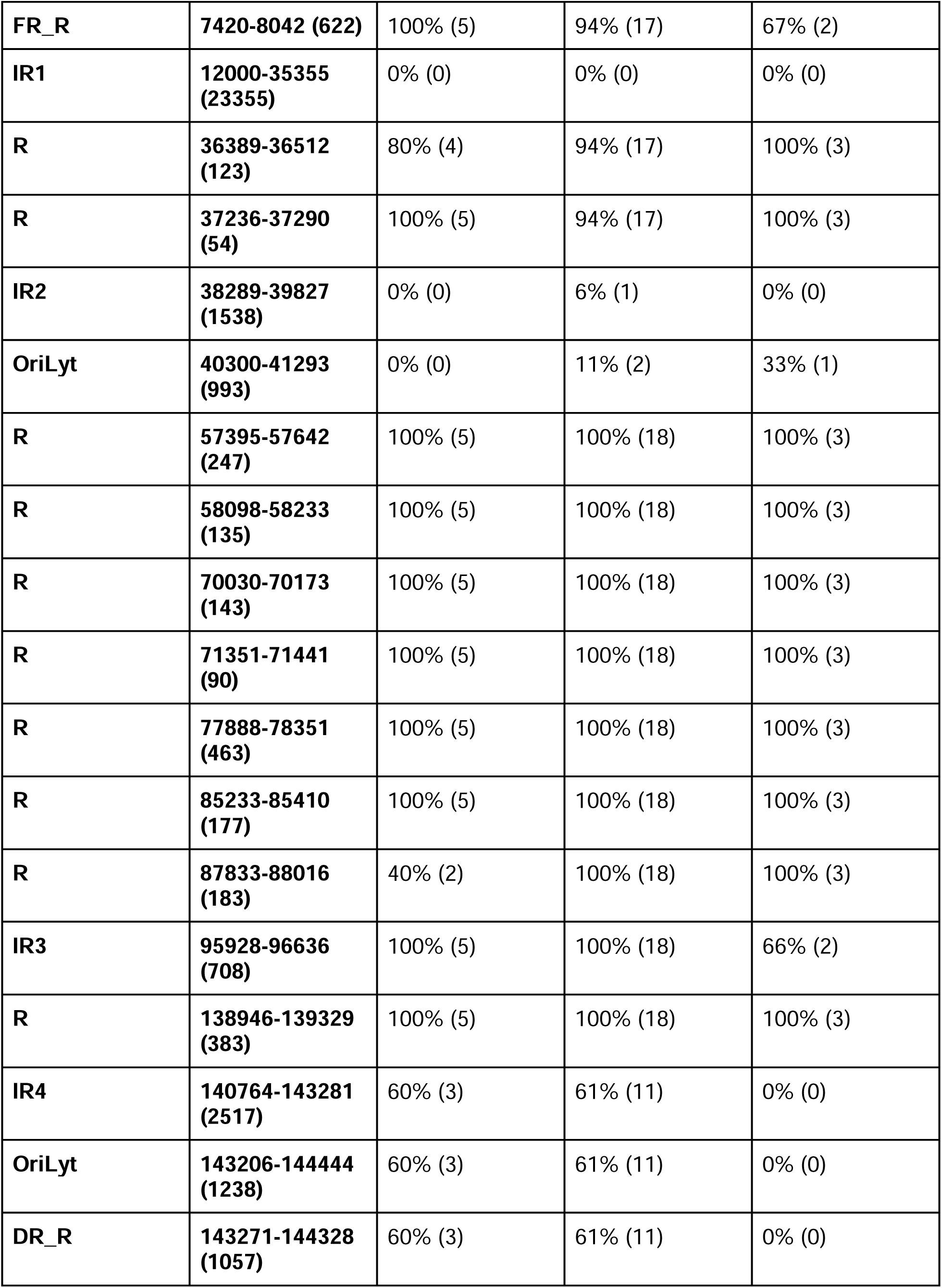
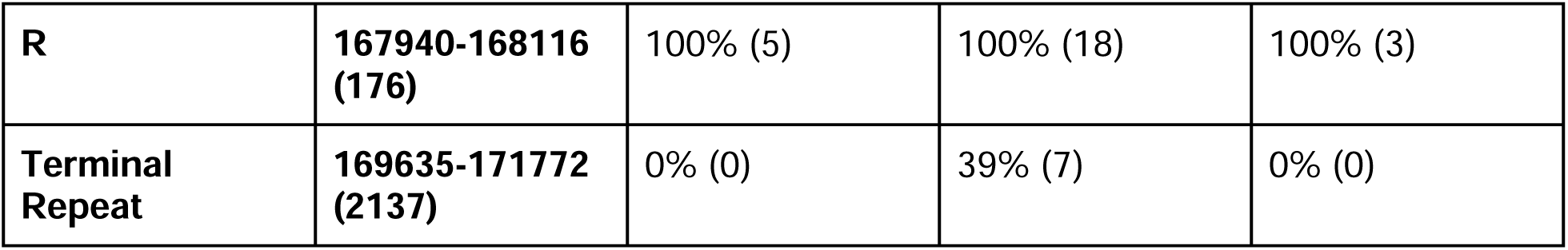
Percentage of Samples with Continuous Assembly Coverage of Repeat Regions.

### ONT assemblies most resembled Illumina counterparts

A phylogenetic tree of all ONT and Illumina assemblies showed that the ONT assemblies most resembled their previous Illumina assemblies counterparts when available (**Figure 5**). As expected, there was a clear separation between assemblies based on EBV type. All ONT assemblies also had shorter or similar branch lengths compared to their Illumina assemblies, which may be indicative of a lower or comparable error rate in the new long-read assemblies, as random errors are often expected to extend branch lengths (**Supplementary** Figures 1-14).^64,65^

**Figure 5.**
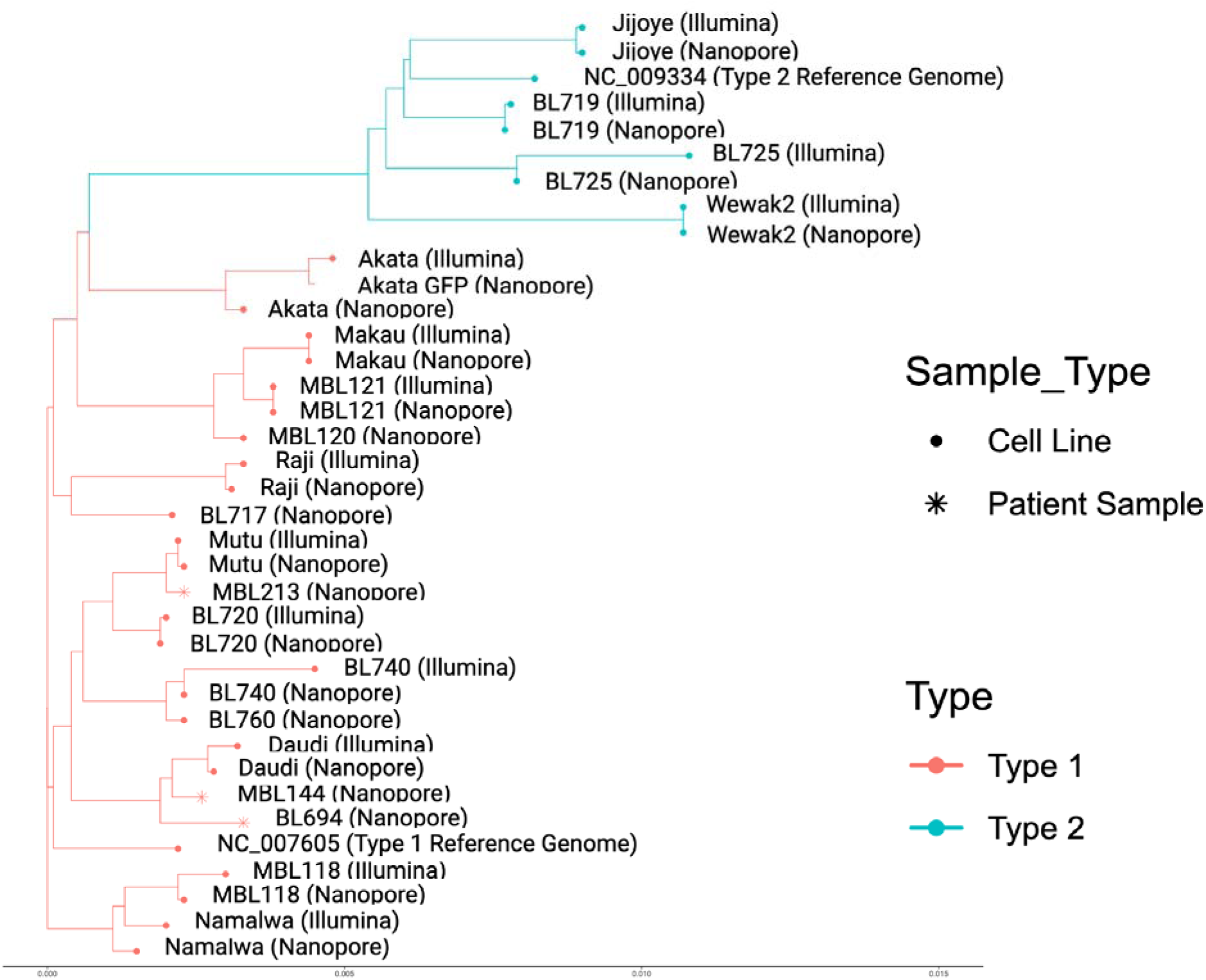
Phylogenetic Analysis. Phylogenetic tree for ONT assemblies of the 18 tumor cell lines and 3 patient tumor samples as well as 14 of their available Illumina assemblies (Akata, Daudi, BL719, BL720, BL725, BL740, Jijoye, Makau, Mutu I, Namalwa, Raji, Wewak2, MBL118, and MBL121).

## DISCUSSION

We developed and optimized a new protocol for selective whole genome amplification combined with targeted PCR enrichment followed by long-read sequencing that significantly enriched the EBV genome in tumor cell lines and patient tumor samples. Across 18 cell lines and three patient tumor biopsies, the resulting assemblies covered 99.62% and 99.68% of the unique genome, respectively, along with full assembly coverage in 14 of the 21 repeat regions for over 80% of the cell lines and 12 of the 21 for all patient tumor samples. They also showed high concordance with Illumina assemblies at 99.61% similarityAlthough it required additional PCR targeting, all of our assemblies covered challenging but biologically significant regions such as *EBNA2*. The overall improvements now allows rapid interrogation of EBV genomes from infected tumors and cell lines scaling from a few to hundreds of genomes.

The amplification pipeline was optimized in many aspects to yield the highest sequencing output and percentage of reads mapping to EBV as well as for scalability and simplicity. For example, despite ONT sequencing requiring a higher amount of input DNA than Illumina, this new pipeline generated enough DNA material (400 ng) per barcode after sWGA, multiplex PCR, and singleplex PCR from only 12.5 ng of initial input DNA.^66^ This pipeline was also optimized for maximum selective amplification with a dNTP concentration of 30/30/5/5 mM for G/C/A/T, likely due to the higher GC content of the EBV genome compared to the human genome.^14,18,67^ Debranching on both the post-sWGA and post-PCR reactions increased sequencing output on the ONT flow cell and by extension, the cost-effectiveness of the pipeline. Furthermore, the gradient annealing temperature of the PCR reaction allowed multiplex primers, which reduced the PCR reaction to a single tube and increased scalability. The combination of our post-sWGA, post-multiplex PCR, and post-singleplex PCR products enabled more uniform amplification of the EBV genome as well.

Compared to prior enrichment methods, this pipeline also significantly enriched the EBV genome in tumor cell lines with as low as 1 EBV copy/cell without expensive library preparation and extensive time. Note that while selected cell lines such as Namalwa containing 1-2 EBV copies/cell were successfully amplified and sequenced, most have at least 20 copies/cell as seen in these 18 selected cell lines.^17,68^ The laboratory infrastructure and costs associated with Illumina sequencing machines diminish its accessibility for laboratories in resource-limited areas such as those in Africa where BL is most prevalent. Current existing methods using the old Phi29™ DNA polymerase take up to 16 hours for sWGA alone. In contrast, this new method did not require RNA or DNA baits, and library preparation for a given sample including sWGA and debranching took approximately five hours although a higher number of samples increases preparation time.

Furthermore, while prior enrichment methods leveraged Illumina sequencing and Phi29 polymerase, both of which tend to drop coverage in GC-rich regions, our percent of ONT bases mapping to EBV after sWGA slightly positively correlated with the GC content of the assemblies, suggesting samples with higher GC content actually experienced marginally higher coverage in this new pipeline. This is likely explained by the improvements in GC bias in ONT sequencing and the new EquiPhi29™ polymerase combined with our provision of relatively more dGTP and dCTP. Studies have also reported the lowest GC bias and highest target yield within 2 hours with the EquiPhi29™ polymerase compared to other commercially available Phi29™ polymerases.^69,70^

The ONT assemblies showed several improvements compared to their Illumina counterparts in accuracy, length, cost, and time. While one of ONT sequencing’s most prominent weaknesses has been its lower accuracy compared to Illumina, this pipeline has produced assemblies that are extremely similar to Illumina at 99.61% accuracy and cover the vast majority of the unique genome including key genes and most smaller repeat regions. ONT base-calling technologies are also rapidly improving with some sequencing kits yielding read accuracy greater than 99%.^71^ Notably, in the phylogenetic tree analysis, the ONT assemblies clustered most closely with their Illumina counterparts. In fact, there is a tradeoff between the error rate and number of single-nucleotide polymorphisms (SNPs).^72^ Thus, since the ONT assemblies have shorter branch lengths compared to Illumina assemblies, they have fewer variants relative to the other EBV genomes, suggesting that there may actually be fewer assembly errors in the ONT assemblies compared to Illumina.

Recent studies have reported that long reads can improve assembly accuracy compared to short reads and generate almost perfect *de novo* assemblies although accuracy may depend on the organism.^73,74^ Assemblies from this pipeline also covered all genes including *EBNA2, EBNA3, BZLF1, EBNA1, BALF5, LMP1*, and *LMP2*, especially with the addition of the singleplex PCR amplification of *EBNA2* and *LMP2*. These genes are known to play critical roles in B-cell growth transformation and tumorigenesis in BL. For example, *EBNA2* is one of the first viral genes expressed in EBV-infected B-cells and serves as a transcriptional regulator of both cellular and viral genes.^75^ LMP1 is an oncoprotein that activates NF-κB,^76^ while *BZLF1* activates lytic gene expression and helps initiate lytic replication.^77^ That this pipeline covered all crucial genes has significant implications for the utility of the pipeline in future EBV studies. Prior viral enrichment methods masked the defined repeat regions and any repeat regions larger than 100 nucleotides in their assemblies. While the EBV genome contains many repeat regions that encode proteins with critical roles in the viral life cycle such as latency, many assemblies are similarly masked at their repeat regions.^78,79^ In contrast, these new ONT assemblies included many of the repeat regions, albeit they cannot traverse the largest repeats such as *IR1*. This is likely due to ONT’s longer read lengths, as most of the repeat regions up to 1000 bps demonstrated continuous assembly coverage across all BL720 dilutions, cell lines, and patient samples, which generally corresponds to the median read lengths. This is important, because the EBV genome contains many repeat regions that encode proteins with critical roles in the viral life cycle such as latency.^78,79^ In this new pipeline, assemblies used as low as 148 megabases of sequencing reads for some cell lines such as Akata. This indicates that in addition to producing an accurate assembly, this method is cost-effective compared to previous methods. ONT’s real-time sequencing ability also means that researchers could sequence a single sample of interest without using up the entire flow cell and save it for future use. While Illumina sequencing has significantly decreased in cost over the last several years, this pipeline makes ONT cheaper for sequencing EBV genomes, especially when taking into account the need for RNA capture baits in Illumina.^14^ Furthermore, ONT sequencing has barcoding kits that enable multiplexing on a large scale. These samples were sequenced with the 24-barcode kit, so eight samples with three barcodes each (sWGA, multiplex PCR, and singleplex PCR) were simultaneously sequenced. The 96-barcoding kit has recently been released for early access, potentially allowing up to 32 samples per sequencing run. Lastly, due to ONT’s capability for real-time data generation, this pipeline could be completed within a day for a sample from initial sWGA to final genome assembly,^80^ further strengthening its future clinical application to quickly sequence and characterize viral variants that may have prognostic significance.

Limitations of this study included the method’s inability to amplify samples with less than 1 EBV copy/cell and traverse the 23kb-long *IR1* repetitive region in these EBV genomes, because while the method produced reads averaging a couple kilobases, which was longer than Illumina’s 200 bp-long reads, its reads were still not long enough for *IR1.* These seem to be a general limitation of Phi29 branch amplification. Regardless, the accuracy, length, cost, and preparation time of this pipeline make it a considerable improvement from prior EBV enrichment methods and especially useful for laboratories in Africa where eBL is most widespread.

## CONCLUSION

This method improves selective amplification of the EBV genome from tumor cell lines and patient tumor biopsies, allowing for direct long-read sequencing on a portable ONT sequencer and accurate assembly of the whole genome at a cost-effective level. The accessibility of this pipeline makes it especially useful for laboratories with limited resources and lays the foundation for rapid viral assessment in Africa and characterization of EBV in tumor cell lines.

## DATA AND RESOURCE AVAILABILITY

All coding scripts used in the pipeline are available on Github at https://github.com/bailey-lab/EBV-SWGA. The custom script for final assembly can be found as jbam_tools_v04.py, and the “--help” command can be used for more information on how to run the commands. All EBV genome sequences have been deposited to NCBI GenBank (accession number PV091352 for strain Jijoye, PV091353 for strain BL719, PV091354 for strain BL725, PV091355 for strain Wewak2, PV091356 for strain Akata, PV091357 for strain Makau, PV091358 for strain MBL121, PV091359 for strain Raji, PV091360 for strain Mutu, PV091361 for strain BL720, PV091362 for strain BL740, PV091363 for strain Daudi, PV091364 for strain MBL118, PV091365 for strain Namalwa, PV091366 for strain Akata-GFP, PV091367 for strain BL717, PV091368 for strain BL760, PV091369 for strain MBL120, PV091370 for strain MBL144, PV091371 for strain MBL213, and PV091372 for strain BL694). All raw reads are located in the Sequence Read Archive (SRA) under the following accession numbers: SAMN46725406 for Jijoye, SAMN46725407 for BL719, SAMN46725408 for BL725, SAMN46725409 for Wewak2, SAMN46725410 for Akata, SAMN46725411 for Makau, SAMN46725412 for Akata-GFP, SAMN46725413 for Daudi, SAMN46725414 for BL717, SAMN46725415 for BL720, SAMN46725416 for BL740, SAMN46725417 for BL760, SAMN46725418 for Mutu, SAMN46725419 for Namalwa, SAMN46725420 for Raji, SAMN46725421 for MBL118, SAMN46725422 for MBL120, SAMN46725423 for MBL121, SAMN46725424 for MBL144, SAMN46725425 for MBL213, and SAMN46725426 for BL694. The samples are registered via BioProject PRJNA1219681 at https://www.ncbi.nlm.nih.gov/bioproject/PRJNA1219681.

## CONFIDENTIALITY STATEMENT

All sample/patient IDs are unknown to anyone outside the research group.

## AUTHOR CONTRIBUTIONS

**Isaac E. Kim, Jr.:** Conception and design of the study, data acquisition and interpretation, and drafting and editing the article

**Cliff Oduor:** Design of the study, data acquisition, and drafting and editing the article

**Abebe Fola:** Data acquisition and drafting and editing the article

**Enrique Puig:** Data acquisition and drafting and editing the article

**Sin Ting Hui:** Data acquisition and drafting and editing the article

**Titus K. Maina:** Data acquisition and drafting and editing the article

**Eddy O. Agwati:** Data acquisition and drafting and editing the article

**Kaleb Zuckerman:** Data acquisition and interpretation, and drafting and editing the article

**Rebecca Crudale:** Data acquisition and drafting and editing the article

**Alec Leonetti:** Data acquisition and drafting and editing the article

**MIcah A. Luftig:** Data acquisition and interpretation and drafting and editing the article

**Ann M. Moormann:** Data acquisition and interpretation and drafting and editing the article

**Jeffrey A. Bailey:** Supervision of the project, conception and design of the study, data acquisition and interpretation, and drafting and editing the article

