## Supplementary Tables 1-10, Supplementary Material 1 for "Comparison of Nanopore with Illumina Whole Genome Assemblies of the Epstein-Barr Virus in Burkitt Lymphoma"

**Supplementary Table 1****. Quantitative PCR Results for sWGA Input Samples**

| **Serial Dilution** | **EBV Copies/Cell According to qPCR** |
| --- | --- |
| 27 copies/cell | 27 (confirmed by ddPCR) |
| 13.5 copies/cell | 16.58 |
| 6.75 copies/cell | 4.09 |
| 3.375 copies/cell | 3.20 |
| 1 copy/cell | 1.08 |
| Namalwa | 1.23 |

**Supplementary Table 2****. Full Patient Characteristics**

| **Sample** | **Sex** | **Age (years)** | **Tumor Site** | **EBV Copies/Cell** | **EBV Type** |
| --- | --- | --- | --- | --- | --- |
| **Tumor Cell Line** |  |  |  |  |  |
| Akata | Female | 1-5 | Abdomen | 5 | 1 |
| Akata GFP (Takada) | Female | 1-5 | Abdomen | 14 | 1 |
| Daudi | Male | 16-20 | Jaw | 7 | 1 |
| BL717 | Female | 6-10 | Orbit | 26 | 1 |
| BL719 | Male | 1-5 | Jaw | 21 | 2 |
| BL720 | Male | 6-10 | Jaw | 28 | 1 |
| BL725 | Male | 1-5 | Abdomen | 20 | 2 |
| BL740 | Female | 6-10 | Jaw | 25 | 1 |
| BL760 | Male | 6-10 | Jaw | 36 | 1 |
| Jijoye | Male | 6-10 | Abdomen | 20 | 2 |
| Makau | N/A | N/A | N/A | 78 | 1 |
| Mutu I | Male | N/A | N/A | 123 | 1 |
| Namalwa | Female | 1-5 | N/A | 1 | 1 |
| Raji | Male | 11-15 | Jaw | 48 | 1 |
| Wewak2 |  |  |  | 132 | 2 |
| MBL118 | Female | 1-5 | Jaw | 92 | 1 |
| MBL120 | Male | 6-10 | Neck | 41 | 1 |
| MBL121 | N/A | N/A | Abdomen | 45 | 1 |
| **Patient Tumor Samples** |  |  |  |  |  |
| MBL144 | F | 6-10 | Abdomen | 36 | 1 |
| MBL213 | M | 6-10 | Abdomen | 42 | 1 |
| BL694 | F | 6-10 | N/A | 8 | 1 |

**Supplementary Table 3****. sWGA optimization results for varied input DNA and dNTP concentrations**

| **Variable** | **BL720 EBV input copies/cell** | **Total DNA input (ng)** | **dNTP Composition in sWGA (G/C/T/A)** | **% Total Reads Mapping to EBV after sWGA** | **EBV/Human Enrichment** |
| --- | --- | --- | --- | --- | --- |
| Total DNA Input | 27 | 100 | 30/30/5/5 | 10.6 | 150.3 |
|  |  | 50 |  | 19.7 | 279.5 |
|  |  | 25 |  | 31.0 | 441.0 |
|  |  | 12.5 |  | 41.0 | 583.1 |
|  |  | 6.25 |  | 36.2 | 514.7 |
|  |  | 1 |  | 4.7 | 67.0 |
|  | 13.5 | 100 |  | 3.8 | 106.7 |
|  |  | 50 |  | 8.7 | 246.0 |
|  |  | 25 |  | 15.1 | 429.2 |
|  |  | 12.5 |  | 22.0 | 624.6 |
|  |  | 6.25 |  | 24.0 | 682.9 |
|  |  | 1 |  | 2.3 | 64.9 |
|  | 6.75 | 100 |  | 2.2 | 125.7 |
|  |  | 50 |  | 4.8 | 273.1 |
|  |  | 25 |  | 8.6 | 490.9 |
|  |  | 12.5 |  | 10.8 | 616.7 |
|  |  | 6.25 |  | 11.6 | 660.5 |
|  |  | 1 |  | 3.3 | 187.7 |
|  | 3.375 | 100 |  | 2.4 | 270.8 |
|  |  | 50 |  | 3.2 | 362.9 |
|  |  | 25 |  | 4.2 | 476.7 |
|  |  | 12.5 |  | 5.2 | 595.0 |
|  |  | 6.25 |  | 4.4 | 504.0 |
|  |  | 1 |  | 2.7 | 310.6 |
|  | 1 | 100 |  | 0.7 | 280.3 |
|  |  | 50 |  | 0.9 | 357.1 |
|  |  | 25 |  | 1.2 | 464.6 |
|  |  | 12.5 |  | 1.8 | 679.7 |
|  |  | 6.25 |  | 0.9 | 349.4 |
|  |  | 1 |  | 1.8 | 683.5 |
| dNTP Composition | 27 | 12.5 | 10-10 mM GC/AT | 18.7 | 266.5 |
|  |  |  | 30-10 mM GC/AT | 60.6 | 861.6 |
|  |  |  | 30-5 mM GC/AT | 61.6 | 875.9 |
|  | 13.5 |  | 10-10 mM GC/AT | 14.3 | 406.7 |
|  |  |  | 30-10 mM GC/AT | 39.7 | 1127.8 |
|  |  |  | 30-5 mM GC/AT | 38.2 | 1086.8 |
|  | 6.75 |  | 10-10 mM GC/AT | 7.2 | 408.5 |
|  |  |  | 30-10 mM GC/AT | 26.7 | 1520.6 |
|  |  |  | 30-5 mM GC/AT | 27.0 | 1537.7 |
|  | 3.375 |  | 10-10 mM GC/AT | 4.5 | 508.6 |
|  |  |  | 30-10 mM GC/AT | 20.5 | 2329.0 |
|  |  |  | 30-5 mM GC/AT | 18.8 | 2136.7 |
|  | 1 |  | 10-10 mM GC/AT | 4.0 | 1543.6 |
|  |  |  | 30-10 mM GC/AT | 13.1 | 5026.4 |
|  |  |  | 30-5 mM GC/AT | 12.7 | 4861.3 |

**Supplementary Table 4****. Percentage of Bases Mapping to EBV and Enrichment Statistics**

| **Sample** | **EBV Copies/Cell** | **% GC Content of Assembly** | **Post-sWGA: % EBV-Mapping Bases (x-fold Enrichment)** | **Post-sWGA: % Human-Mapping Bases (x-fold Enrichment)** | **Post-Multiplex PCR: % EBV-Mapping Bases (x-fold Enrichment)** | **Post-Multiplex PCR: % Human-Mapping Bases (x-fold Enrichment)** | **Post-Singleplex PCR: % EBV-Mapping Bases (x-fold Enrichment)** | **Post-Singleplex PCR: % Human-Mapping Bases (x-fold Enrichment)** |
| --- | --- | --- | --- | --- | --- | --- | --- | --- |
| **BL720 Dilution** |  |  |  |  |  |  |  |  |
| 27 copies/cell | 27 | 48.5 | 46.4 (660x) | 64.7 (0.7x) | 93.3 (1328x) | 19.4 (0.2x) | N/A | N/A |
| 13.5 copies/cell | 13.5 | 47.1 | 25.9 (736x) | 81.3 (0.8x) | 81.9 (2331x) | 32.3 (0.3x) | N/A | N/A |
| 6.75 copies/cell | 6.75 | 47.8 | 17.6 (1002x) | 87.4 (0.9x) | 74.0 (4211x) | 40.5 (0.4x) | N/A | N/A |
| 3.375 copies/cell | 3.375 | 46.6 | 12.0 (1370x) | 88.8 (0.9x) | 62.2 (7072x) | 51.3 (0.5x) | N/A | N/A |
| 1 copy/cell | 1 | 46.0 | 8.6 (3283x) | 92.4 (0.9x) | 47.2 (18142x) | 64.7 (0.7x) | N/A | N/A |
| **Tumor Cell Line** |  |  |  |  |  |  |  |  |
| Akata | 5 | 46.1 | 8.8 (675x) | 97.6 (1.0x) | 21.0 (1614x) | 95.3 (1.0x) | 12.4 (950x) | 98.2 (1.0x) |
| Akata GFP (Takada) | 14 | 46.9 | 15.7 (431x) | 91.0 (0.9x) | 83.2 (2283x) | 38.1 (0.4x) | 58.5 (1606x) | 68.0 (0.6x) |
| Daudi | 7 | 46.6 | 15.4 (842x) | 91.7 (0.9x) | 23.3 (1279x) | 94.2 (0.9x) | 48.8 (2675x) | 72.1 (0.7x) |
| BL717 | 26 | 48.8 | 40.0 (591x) | 68.2 (0.7x) | 91.2 (1348x) | 20.7 (0.2x) | 81.6 (1206x) | 42.0 (0.4x) |
| BL719 | 21 | 48.4 | 37.6 (687x) | 70.9 (0.7x) | 72.5 (1326x) | 70.2 (0.7x) | 74.5 (1364x) | 44.7 (0.5x) |
| BL720 | 28 | 48.0 | 46.4 (637x) | 64.7 (0.7x) | 79.8 (1095x) | 32.0 (0.3x) | 51.3 (704x) | 68.0 (0.7x) |
| BL725 | 20 | 49.1 | 33.0 (634x) | 74.8 (0.8x) | 91.2 (1751x) | 21.9 (0.2x) | 66.6 (1280x) | 53.4 (0.5x) |
| BL740 | 25 | 48.4 | 49.5 (760x) | 57.5 (0.6x) | 95.4 (1466x) | 14.8 (0.2x) | 79.4 (1220x) | 39.6 (0.4x) |
| BL760 | 36 | 48.9 | 59.1 (631x) | 49.5 (0.5x) | 95.6 (1021x) | 16.0 (0.2x) | 81.7 (872x) | 37.2 (0.4x) |
| Jijoye | 20 | 48.8 | 25.7 (493x) | 79.9 (0.8x) | 90.6 (1741x) | 23.0 (0.2x) | 63.3 (1217x) | 56.9 (0.6x) |
| Makau | 78 | 49.6 | 44.2 (218x) | 63.8 (0.6x) | 92.8 (458x) | 19.7 (0.2x) | 76.2 (375.9x) | 44.9 (0.5x) |
| Mutu I | 123 | 47.6 | 65.0 (204x) | 44.1 (0.4x) | 96.6 (303x) | 13.1 (0.1x) | 83.8 (262x) | 34.4 (0.4x) |
| Namalwa | 1 | 48.1 | 8.6 (3307x) | 92.8 (0.9x) | 15.8 (6051x) | 96.0 (1.0x) | 19.2 (7354x) | 80.5 (0.8x) |
| Raji | 48 | 47.1 | 41.2 (330x) | 65.0 (0.7x) | 92.3 (740x) | 20.1 (0.2x) | 75.7 (607x) | 46.4 (0.5x) |
| Wewak2 | 132 | 48.9 | 53.9 (157x) | 52.1 (0.5x) | 96.5 (282x) | 13.1 (0.1x) | 84.0 (245x) | 33.8 (0.3x) |
| MBL118 | 92 | 49.4 | 45.1 (189x) | 61.3 (0.6x) | 45.1 (189x) | 61.3 (0.6x) | 68.8 (288x) | 52.6 (0.5x) |
| MBL120 | 41 | 48.8 | 35.8 (336x) | 70.5 (0.7x) | 35.8 (336x) | 70.5 (0.7x) | 58.4 (547x) | 59.5 (0.6x) |
| MBL121 | 45 | 48.0 | 27.3 (233x) | 78.1 (0.8x) | 27.3 (233x) | 78.1 (0.8x) | 46.0 (393x) | 70.6 (0.7x) |
| **Patient Tumor Samples** |  |  |  |  |  |  |  |  |
| MBL144 | 36 | 47.2 | 30.7 (328x) | 79.3 (0.8x) | 78.7 (840x) | 37.5 (0.4x) | 48.8 (521x) | 65.4 (0.7x) |
| MBL213 | 42 | 47.0 | 30.6 (280x) | 80.3 (0.8x) | 66.8 (612x) | 52.9 (0.5x) | 41.8 (382.8x) | 78.5 (0.8x) |
| BL694 | 8 | 47.1 | 17.9 (861x) | 90.4 (0.9x) | 57.1 (2740x) | 57.4 (0.6x) | 49.8 (2389x) | 72.1 (0.7x) |

**Supplementary Table 5****. Median Read Lengths of EBV-Mapping Reads**

| **Sample** | **Post-sWGA (bp)** | **Post-Multiplex PCR (bp)** | **Post-Singleplex PCR (bp)** |
| --- | --- | --- | --- |
| **BL720 Dilution** |  |  |  |
| 27 copies/cell | 1544 | 1272 | N/A |
| 13.5 copies/cell | 1538 | 1258 | N/A |
| 6.75 copies/cell | 1413 | 1233 | N/A |
| 3.375 copies/cell | 1449 | 1119 | N/A |
| 1 copy/cell | 1382 | 915 | N/A |
| **Tumor Cell Line** |  |  |  |
| Akata | 1535 | 938 | 1030 |
| Akata GFP (Takada) | 1432 | 1532 | 1274 |
| Daudi | 1562 | 706 | 1249 |
| BL717 | 1657 | 1473 | 1737 |
| BL719 | 1625 | 894 | 1698 |
| BL720 | 1544 | 1359 | 1731 |
| BL725 | 1776 | 1435 | 1756 |
| BL740 | 1905 | 1448 | 1719 |
| BL760 | 1942 | 1455 | 1656 |
| Jijoye | 1936 | 1430 | 1750 |
| Makau | 2141 | 1450 | 1366 |
| Mutu I | 1714 | 1438 | 1227 |
| Namalwa | 1720 | 816 | 1497 |
| Raji | 2067 | 1422 | 1468 |
| Wewak2 | 1814 | 1417 | 1447 |
| MBL118 | 1938 | 1938 | 1509 |
| MBL120 | 1951 | 1951 | 1292 |
| MBL121 | 1924 | 1924 | 1167 |
| **Patient Tumor Samples** |  |  |  |
| MBL144 | 1839 | 1402 | 1503 |
| MBL213 | 2366 | 1432 | 1455 |
| BL694 | 1672 | 1102 | 1593 |

**Supplementary Table 6****. Median Read Depth Coverage of Repeat Regions (First Half)**

| **Repeat Region** | **OriP** | **FR_R** | **IR1** | **R** | **R** | **IR2** | **OriLyt** | **R** | **R** | **R** | **R** |
| --- | --- | --- | --- | --- | --- | --- | --- | --- | --- | --- | --- |
| **Position on Reference Genome NC_007605** | **7314-9312** | **7420-8042** | **12000-35355** | **36389-36512** | **37236-37290** | **38289-39827** | **40300-41293** | **57395-57642** | **58098-58233** | **70030-70173** | **71351-71441** |
| **Length** | **1998** | **622** | **23355** | **123** | **54** | **1538** | **993** | **247** | **135** | **143** | **90** |
| **BL720 Dilution** |  |  |  |  |  |  |  |  |  |  |  |
| 27 copies/cell | 131 | 120 | 0 | 39 | 66 | 4 | 3,517 | 524 | 308 | 2,086 | 1,996 |
| 13.5 copies/cell | 217 | 184 | 0 | 32 | 129 | 1 | 8,411 | 1,192 | 933 | 320 | 4,022 |
| 6.75 copies/cell | 67 | 60 | 0 | 30 | 77 | 0 | 6,923 | 1,686 | 1,614 | 236 | 3,329 |
| 3.375 copies/cell | 32 | 33 | 0 | 10 | 61 | 0 | 6,596 | 530 | 328 | 117 | 2,445 |
| 1 copy/cell | 559 | 558 | 0 | 6 | 32 | 0 | 2,733 | 371 | 271 | 53 | 688 |
| **Tumor Cell Line** |  |  |  |  |  |  |  |  |  |  |  |
| Akata | 15 | 9 | 0 | 42 | 35 | 1 | 1,739 | 213 | 79 | 55 | 266 |
| Akata GFP (Takada) | 6 | 2 | 0 | 25 | 35 | 1 | 224 | 48 | 39 | 94 | 335 |
| Daudi | 106 | 97 | 0 | 14 | 14 | 0 | 4,793 | 193 | 73 | 92 | 407 |
| BL717 | 38 | 29 | 0 | 25 | 32 | 1 | 368 | 177 | 300 | 198 | 808 |
| BL719 | 65 | 62 | 1 | 16 | 120 | 4 | 8,072 | 2,042 | 933 | 381 | 1,571 |
| BL720 | 189 | 178 | 0 | 129 | 191 | 5 | 4,197 | 847 | 944 | 2,537 | 2,775 |
| BL725 | 43 | 18 | 0 | 23 | 117 | 1 | 269 | 142 | 176 | 119 | 448 |
| BL740 | 110 | 27 | 0 | 324 | 307 | 1 | 426 | 228 | 361 | 120 | 1,342 |
| BL760 | 65 | 30 | 0 | 130 | 112 | 2 | 236 | 222 | 369 | 186 | 455 |
| Jijoye | 21 | 20 | 0 | 20 | 62 | 0 | 275 | 99 | 59 | 89 | 286 |
| Makau | 33 | 27 | 0 | 312 | 276 | 2 | 246 | 332 | 416 | 222 | 406 |
| Mutu I | 81 | 44 | 0 | 899 | 711 | 2 | 254 | 268 | 564 | 478 | 528 |
| Namalwa | 18 | 15 | 0 | 194 | 100 | 0 | 1,049 | 1,021 | 143 | 97 | 375 |
| Raji | 75 | 21 | 0 | 377 | 328 | 1 | 412 | 213 | 231 | 325 | 567 |
| Wewak2 | 134 | 105 | 1 | 66 | 521 | 0 | 226 | 383 | 69 | 293 | 949 |
| MBL118 | 86 | 24 | 0 | 552 | 480 | 1 | 212 | 51 | 52 | 207 | 476 |
| MBL120 | 28 | 26 | 0 | 290 | 275 | 1 | 410 | 89 | 87 | 310 | 691 |
| MBL121 | 33 | 28 | 0 | 365 | 265 | 1 | 437 | 56 | 23 | 99 | 374 |
| **Patient Tumor Samples** |  |  |  |  |  |  |  |  |  |  |  |
| MBL144 | 116 | 436 | 0 | 130 | 184 | 0 | 7,779 | 856 | 561 | 404 | 4,321 |
| MBL213 | 276 | 2,493 | 0 | 49 | 39 | 0 | 7,304 | 751 | 557 | 291 | 3,636 |
| BL694 | 24 | 1,139 | 0 | 338 | 337 | 0 | 4,485 | 135 | 80 | 156 | 940 |

**Supplementary Table 7****. Median Read Depth Coverage of Repeat Regions (Second Half)**

| **Repeat Region** | **R** | **R** | **R** | **IR3** | **R** | **IR4** | **OriLyt** | **DR_R** | **R** | **Terminal Repeat** |
| --- | --- | --- | --- | --- | --- | --- | --- | --- | --- | --- |
| **Position on Reference Genome NC_007605** | **77888-78351** | **85233-85410** | **87833-88016** | **95928-96636** | **138946-139329** | **140764-143281** | **143206-144444** | **143271-144328** | **167940-168116** | **169635-171772** |
| **Length** | **463** | **177** | **183** | **708** | **383** | **2517** | **1238** | **1057** | **176** | **2137** |
| **BL720 Dilution** |  |  |  |  |  |  |  |  |  |  |
| 27 copies/cell | 435 | 366 | 6 | 54 | 801 | 18 | 2,776 | 2,762 | 1,019 | 3 |
| 13.5 copies/cell | 943 | 640 | 10 | 122 | 1,905 | 13 | 7,229 | 7,190 | 1,698 | 4 |
| 6.75 copies/cell | 810 | 580 | 13 | 116 | 1,607 | 4 | 5,406 | 5,369 | 1,234 | 3 |
| 3.375 copies/cell | 366 | 258 | 8 | 65 | 979 | 0 | 4,007 | 3,981 | 915 | 3 |
| 1 copy/cell | 75 | 77 | 5 | 15 | 534 | 2 | 1,481 | 1,471 | 351 | 0 |
| **Tumor Cell Line** |  |  |  |  |  |  |  |  |  |  |
| Akata | 29 | 51 | 70 | 63 | 534 | 2 | 1,118 | 1,098 | 271 | 5 |
| Akata GFP (Takada) | 35 | 14 | 95 | 19 | 377 | 11 | 490 | 480 | 167 | 16 |
| Daudi | 350 | 673 | 71 | 265 | 888 | 2 | 3,231 | 3,206 | 345 | 14 |
| BL717 | 64 | 46 | 130 | 66 | 640 | 16 | 526 | 517 | 376 | 15 |
| BL719 | 92 | 412 | 209 | 235 | 2,813 | 22 | 3,587 | 3,558 | 3,685 | 13 |
| BL720 | 572 | 399 | 55 | 108 | 1,065 | 26 | 3,280 | 3,256 | 1,452 | 23 |
| BL725 | 124 | 59 | 175 | 108 | 817 | 36 | 943 | 932 | 441 | 203 |
| BL740 | 76 | 32 | 414 | 82 | 569 | 20 | 661 | 651 | 490 | 206 |
| BL760 | 67 | 34 | 30 | 36 | 583 | 46 | 485 | 478 | 313 | 83 |
| Jijoye | 62 | 40 | 164 | 155 | 750 | 26 | 887 | 870 | 333 | 104 |
| Makau | 41 | 38 | 353 | 46 | 301 | 51 | 801 | 790 | 465 | 75 |
| Mutu I | 67 | 43 | 257 | 97 | 426 | 12 | 320 | 316 | 290 | 189 |
| Namalwa | 36 | 124 | 184 | 30 | 189 | 0 | 323 | 320 | 152 | 26 |
| Raji | 62 | 565 | 33 | 135 | 149 | 3 | 309 | 306 | 87 | 117 |
| Wewak2 | 70 | 30 | 148 | 83 | 1,027 | 10 | 524 | 518 | 533 | 511 |
| MBL118 | 30 | 32 | 392 | 107 | 483 | 20 | 504 | 500 | 241 | 96 |
| MBL120 | 46 | 38 | 359 | 83 | 250 | 6 | 375 | 370 | 306 | 186 |
| MBL121 | 53 | 37 | 357 | 49 | 214 | 2 | 374 | 369 | 347 | 52 |
| **Patient Tumor Samples** |  |  |  |  |  |  |  |  |  |  |
| MBL144 | 239 | 313 | 109 | 15 | 1,257 | 0 | 4,832 | 4,810 | 1,372 | 4 |
| MBL213 | 28 | 207 | 1,431 | 1 | 239 | 0 | 5,110 | 5,098 | 398 | 0 |
| BL694 | 140 | 74 | 341 | 48 | 134 | 0 | 1,923 | 1,912 | 518 | 7 |

**Supplementary Table 8****. Assembly Statistics**

| **Sample** | **Assembly Breadth of Coverage (without repeats)** | **Assembly Similarity to Illumina** | **N50 (L50)** |
| --- | --- | --- | --- |
| **BL720 Copies/Cell** |  |  |  |
| 27 | 98.60% | 99.86% | 168145 (1) |
| 13.5 | 97.04% | 99.94% | 166725 (1) |
| 6.75 | 96.62% | 99.92% | 164238 (1) |
| 3.375 | 95.19% | 99.94% | 166145 (1) |
| 1 | 93.90% | 99.91% | 165043 (1) |
| **Tumor Cell Line** |  |  |  |
| Akata | 98.67% | 99.47% | 171484 (1) |
| Akata GFP (Takada) | N/A | N/A | 169204 (1) |
| Daudi | 99.82 | 99.69% | 169464 (1) |
| BL717 | N/A | N/A | 168962 (1) |
| BL719 | 99.68 | 99.85% | 168962 (1) |
| BL720 | 99.54 | 99.77% | 171498 (1) |
| BL725 | 99.85 | 99.95% | 172010 (1) |
| BL740 | 100.00 | 99.90% | 168625 (1) |
| BL760 | N/A | N/A | 171674 (1) |
| Jijoye | 100.00 | 99.49% | 171768 (1) |
| Makau | 100.00 | 99.56% | 168864 (1) |
| Mutu I | 98.68 | 99.93% | 171582 (1) |
| Namalwa | 100.00 | 98.36% | 168779 (1) |
| Raji | 100.00 | 99.19% | 169286 (1) |
| Wewak2 | 100.00 | 99.78% | 172215 (1) |
| MBL118 | 99.56 | 99.85% | 171603 (1) |
| MBL120 | N/A | N/A | 171333 (1) |
| MBL121 | 98.95 | 99.73% | 167320 (1) |
| **Patient Tumor Samples** |  |  |  |
| MBL144 | 98.72% | N/A | 169177 (1) |
| MBL213 | 98.71% | N/A | 167419 (1) |
| BL694 | 98.60% | N/A | 169422 (1) |

**Supplementary Table 9****. Complete Continuous Spanning/Contiguous Assembly Coverage of Repeat Regions (First Half)**

| **Repeat Region** | **OriP** | **FR_R** | **IR1** | **R** | **R** | **IR2** | **OriLyt** | **R** | **R** | **R** | **R** |
| --- | --- | --- | --- | --- | --- | --- | --- | --- | --- | --- | --- |
| **Position on Reference Genome NC_007605** | **7314-9312** | **7420-8042** | **12000-35355** | **36389-36512** | **37236-37290** | **38289-39827** | **40300-41293** | **57395-57642** | **58098-58233** | **70030-70173** | **71351-71441** |
| **Length** | **1998** | **622** | **23355** | **123** | **54** | **1538** | **993** | **247** | **135** | **143** | **90** |
| **BL720 Dilution** |  |  |  |  |  |  |  |  |  |  |  |
| 27 copies/cell | Yes | Yes | No | Yes | Yes | No | No | Yes | Yes | Yes | Yes |
| 13.5 copies/cell | Yes | Yes | No | Yes | Yes | No | No | Yes | Yes | Yes | Yes |
| 6.75 copies/cell | Yes | Yes | No | Yes | Yes | No | No | Yes | Yes | Yes | Yes |
| 3.375 copies/cell | Yes | Yes | No | Yes | Yes | No | No | Yes | Yes | Yes | Yes |
| 1 copy/cell | Yes | Yes | No | No | Yes | No | No | Yes | Yes | Yes | Yes |
| **Tumor Cell Line** |  |  |  |  |  |  |  |  |  |  |  |
| Akata | Yes | Yes | No | Yes | Yes | No | No | Yes | Yes | Yes | Yes |
| Akata GFP (Takada) | No | No | No | Yes | Yes | No | No | Yes | Yes | Yes | Yes |
| Daudi | Yes | Yes | No | No | No | No | No | Yes | Yes | Yes | Yes |
| BL717 | Yes | Yes | No | Yes | Yes | No | No | Yes | Yes | Yes | Yes |
| BL719 | Yes | Yes | No | Yes | Yes | No | No | Yes | Yes | Yes | Yes |
| BL720 | Yes | Yes | No | Yes | Yes | Yes | Yes | Yes | Yes | Yes | Yes |
| BL725 | Yes | Yes | No | Yes | Yes | No | No | Yes | Yes | Yes | Yes |
| BL740 | Yes | Yes | No | Yes | Yes | No | No | Yes | Yes | Yes | Yes |
| BL760 | Yes | Yes | No | Yes | Yes | No | No | Yes | Yes | Yes | Yes |
| Jijoye | Yes | Yes | No | Yes | Yes | No | No | Yes | Yes | Yes | Yes |
| Makau | Yes | Yes | No | Yes | Yes | No | Yes | Yes | Yes | Yes | Yes |
| Mutu I | Yes | Yes | No | Yes | Yes | No | No | Yes | Yes | Yes | Yes |
| Namalwa | Yes | Yes | No | Yes | Yes | No | No | Yes | Yes | Yes | Yes |
| Raji | Yes | Yes | No | Yes | Yes | No | No | Yes | Yes | Yes | Yes |
| Wewak2 | Yes | Yes | No | Yes | Yes | No | No | Yes | Yes | Yes | Yes |
| MBL118 | Yes | Yes | No | Yes | Yes | No | No | Yes | Yes | Yes | Yes |
| MBL120 | Yes | Yes | No | Yes | Yes | No | No | Yes | Yes | Yes | Yes |
| MBL121 | Yes | Yes | No | Yes | Yes | No | No | Yes | Yes | Yes | Yes |
| **Patient Tumor Samples** |  |  |  |  |  |  |  |  |  |  |  |
| MBL144 | Yes | No | No | Yes | Yes | No | No | Yes | Yes | Yes | Yes |
| MBL213 | Yes | Yes | No | Yes | Yes | No | Yes | Yes | Yes | Yes | Yes |
| BL694 | Yes | Yes | No | Yes | Yes | No | No | Yes | Yes | Yes | Yes |

**Supplementary Table 10****. Median Continuous Assembly Coverage of Repeat Regions (Second Half)**

| **Repeat Region** | **R** | **R** | **R** | **IR3** | **R** | **IR4** | **OriLyt** | **DR_R** | **R** | **Terminal Repeat** |
| --- | --- | --- | --- | --- | --- | --- | --- | --- | --- | --- |
| **Position on Reference Genome NC_007605** | **77888-78351** | **85233-85410** | **87833-88016** | **95928-96636** | **138946-139329** | **140764-143281** | **143206-144444** | **143271-144328** | **167940-168116** | **169635-171772** |
| **Length** | **463** | **177** | **183** | **708** | **383** | **2517** | **1238** | **1057** | **176** | **2137** |
| **BL720 Dilution** |  |  |  |  |  |  |  |  |  |  |
| 27 copies/cell | Yes | Yes | Yes | Yes | Yes | Yes | Yes | Yes | Yes | No |
| 13.5 copies/cell | Yes | Yes | No | Yes | Yes | Yes | Yes | Yes | Yes | No |
| 6.75 copies/cell | Yes | Yes | Yes | Yes | Yes | Yes | Yes | Yes | Yes | No |
| 3.375 copies/cell | Yes | Yes | No | Yes | Yes | No | No | No | Yes | No |
| 1 copy/cell | Yes | Yes | No | Yes | Yes | No | No | No | Yes | No |
| **Tumor Cell Line** |  |  |  |  |  |  |  |  |  |  |
| Akata | Yes | Yes | Yes | Yes | Yes | No | No | No | Yes | No |
| Akata GFP (Takada) | Yes | Yes | Yes | Yes | Yes | No | No | No | Yes | Yes |
| Daudi | Yes | Yes | Yes | Yes | Yes | No | No | No | Yes | No |
| BL717 | Yes | Yes | Yes | Yes | Yes | Yes | Yes | Yes | Yes | Yes |
| BL719 | Yes | Yes | Yes | Yes | Yes | Yes | Yes | Yes | Yes | No |
| BL720 | Yes | Yes | Yes | Yes | Yes | No | No | No | Yes | No |
| BL725 | Yes | Yes | Yes | Yes | Yes | Yes | Yes | Yes | Yes | No |
| BL740 | Yes | Yes | Yes | Yes | Yes | Yes | Yes | Yes | Yes | Yes |
| BL760 | Yes | Yes | Yes | Yes | Yes | Yes | Yes | Yes | Yes | Yes |
| Jijoye | Yes | Yes | Yes | Yes | Yes | Yes | Yes | Yes | Yes | No |
| Makau | Yes | Yes | Yes | Yes | Yes | Yes | Yes | Yes | Yes | Yes |
| Mutu I | Yes | Yes | Yes | Yes | Yes | No | No | No | Yes | Yes |
| Namalwa | Yes | Yes | Yes | Yes | Yes | No | No | No | Yes | No |
| Raji | Yes | Yes | Yes | Yes | Yes | No | No | No | Yes | No |
| Wewak2 | Yes | Yes | Yes | Yes | Yes | Yes | Yes | Yes | Yes | No |
| MBL118 | Yes | Yes | Yes | Yes | Yes | Yes | Yes | Yes | Yes | No |
| MBL120 | Yes | Yes | Yes | Yes | Yes | Yes | Yes | Yes | Yes | Yes |
| MBL121 | Yes | Yes | Yes | Yes | Yes | Yes | Yes | Yes | Yes | No |
| **Patient Tumor Samples** |  |  |  |  |  |  |  |  |  |  |
| MBL144 | Yes | Yes | Yes | No | Yes | No | No | No | Yes | No |
| MBL213 | Yes | Yes | Yes | Yes | Yes | No | No | No | Yes | No |
| BL694 | Yes | Yes | Yes | Yes | Yes | No | No | No | Yes | No |

**Supplemental Material 1****. Full Protocol from sWGA to ONT Sequencing**

**Part I: sWGA:**

1. Prepare the following denature mixture for each sample, making sure to pipette mix with each component^1^:

| **Reagent** | **Volume (uL)** | **Final Concentration** |
| --- | --- | --- |
| DNA sample | 12.5 ng |  |
| 100 mM sWGA primer mix* | 3 uL | 10 mM total |
| 10x Reaction Buffer for EquiPhi29 | 1 uL | 1X |
| Nuclease-free water | Up to 10 uL total |  |
| **Total of Denature Mixture** | **10 uL** |  |

* Combine equal volumes of 10 mM sWGA primers listed in Supplementary Table 1 to make a 100 mM sWGA primer mix

2. Heat at 95℃ for 3 minutes, then immediately place on ice for another 5 minutes.
3. Prepare the following sWGA mixture for each sample, making sure to pipette mix with each component:

| **Reagent** | **Volume (uL)** | **Final Concentration** |
| --- | --- | --- |
| Denature Mixture | 10 uL |  |
| 10x Reaction Buffer for EquiPhi29 | 2 | 1X |
| DTT (100 mM) | 0.2 | 1 mM |
| dNTP Mix* | 2 | 3 mM of dGTP, 3 mM of dCTP, 0.5 mM of dATP, and 0.5 mM of dTTP |
| EquiPhi29 DNA Polymerase (10 U/uL) | 1 uL | 10 U |
| Nuclease-free water | 4.8 uL |  |
| **Total of sWGA Mixture** | **20 uL** |  |

* dNTP Mix should be composed of 30 mM of dGTP, 30 mM of dCTP, 5 mM of dATP, and 5 mM of dTTP

4. Incubate at 45℃ for 3 hours, then inactivate polymerase at 65℃ for 10 minutes.
5. Qubit the final sWGA Mixture

^1^ Adapted from ThermoScientific’s EquiPhi29 DNA Polymerase protocol

**Part II: Debranching 1**^2^**:**

6. Prepare the following debranching mixture for each sample, making sure to pipette mix with each component:

| **Reagent** | **Volume** |
| --- | --- |
| 1.5 ug of amplified DNA | x uL |
| NEBuffer 2 | 3 uL |
| T7 Endonuclease I | 1.5 uL |
| Nuclease-free water | Up to 30 uL total |
| **Total of Debranching Mixture** | **30 uL** |

7. Incubate reaction at 37℃ for 15 minutes.
8. Resuspend the AMPure XP beads by vortexing.
9. Prepare the following custom buffer:

| **Reagent** | **Volume** |
| --- | --- |
| 1 M Tris-HCl | 20 uL |
| 0.5 M EDTA pH 8 | 4 uL |
| 5 M NaCl | 640 uL |
| PEG 8000 | 440 uL |
| Nuclease-free water | 888 uL |
| **Total of Custom Buffer** | **1992 uL** |

10. Transfer two aliquots of 1 mL each mixed Agencourt AMPure XP beads into two 1.5 mL Eppendorf DNA LoBind tubes.
11. Pellet the beads on a magnet. Keeping the tube on the magnet, pipette off the supernatant.
12. Wash the beads with 1 mL of nuclease-free water by resuspending the pellet. Return the tube to the magnetic rack, allow the beads to pellet, remove the water using a pipette, and discard.
13. Repeat Step 12.
14. Spin down, and place the tube back on the magnet. Pipette of the residual water.
15. Pool the two bead pellets together by resuspending them in 200 μl of Custom buffer. Then transfer the beads into the remaining Custom buffer.
16. Make up the amplified DNA sample to a total volume of 50 μl in TE buffer, pH 8.
17. Add 90 μl of the custom bead suspension with beads to the DNA sample, and mix by flicking the tube.
18. Thoroughly mix by hand 20 minutes at room temperature, making sure that the solution is not stuck to the tube.
19. Prepare 500 μl of fresh 80% ethanol in nuclease-free water.
20. Spin down the sample and pellet on a magnet until the supernatant is clear and colorless. Keep the tube on the magnet, and pipette off the supernatant.
21. Keep the tube on the magnet and wash the beads with 200 μl of freshly prepared 80% ethanol without disturbing the pellet. Remove the ethanol using a pipette and discard.
22. Repeat the previous step.
23. Spin down and place the tube back on the magnet. Pipette off any residual supernatant. Allow to dry for ~30 seconds, but do not dry the pellet to the point of cracking.
24. Remove the tube from the magnetic rack and resuspend the pellet in 13 μl nuclease-free water.
25. Incubate for 1 minute at 50°C, and then for 5 min at room temperature.
26. Pellet the beads on a magnet until the eluate is clear and colorless.
27. Remove and retain 13 μl of the debranched DNA eluate into a clean 1.5 ml Eppendorf DNA LoBind tube.
28. Qubit the eluate.

^2^ Adapted from Oxford Nanopore Technologies’ Premium Whole Genome Amplification SQK-LSK112 protocol

**Part III: Post-sWGA PCR:**

29. Prepare the following PCR Pool 1 mixture using the PCR Pool 1 primers listed in Supplementary Table 1.

| **Reagent** | **Volume (uL)** |
| --- | --- |
| Pool 1 Primers (36 primers total) | *y* uL each |
| Nuclease-free water | 36*9**y* uL |
| **Total of Primer Pool 1** | 360**y* |

30. Prepare the following PCR Pool 2 mixture using the PCR Pool 2 primers listed in Supplementary Table 1.

| **Reagent** | **Volume (uL)** |
| --- | --- |
| Pool 1 Primers (33 primers total) | *z* uL each |
| Nuclease-free water | 33*9**z* uL |
| **Total of Primer Pool 2** | 330**z* |

31. Prepare the following PCR mixture with two pools, making sure to pipette mix with each component:

| **Reagent** | **Volume (uL)** | **Final Concentration** |
| --- | --- | --- |
| Debranched DNA Eluate | 100 ng |  |
| 2X KAPA HiFi HotStart Ready Mix | 12.5 uL | 1X |
| PCR Primer Mix Pool 1 or 2 10 uM each* | 1.5 uL |  |
| Nuclease-free water | Up to 25 uL total |  |
| **Total of Denature Mixture** | **25 uL** |  |

32. Perform the PCR with the following cycling protocol:

| **Step** | **Temperature** | **Duration** | **Cycles** |
| --- | --- | --- | --- |
| Initial Denaturation | 95°C | 3 min | 1 |
| Denaturation | 98°C | 3 min | 25 |
| Gradient Annealing | 58-49°C | 15 seconds at each °C |  |
| Extension | 72°C | 7 min |  |
| Final Extension | 72°C | 10 min | 1 |

33. Mix the two reaction solutions well.
34. Qubit the final, combined PCR mixture.

**Part IV: Debranching 2:**

35. Repeat Steps 6-28.

**Part V. ONT sequencing (Native Barcoding Kit 24 V14 protocol)**

**Supplementary Figure 1. Jijoye Phylogenetic Tree**

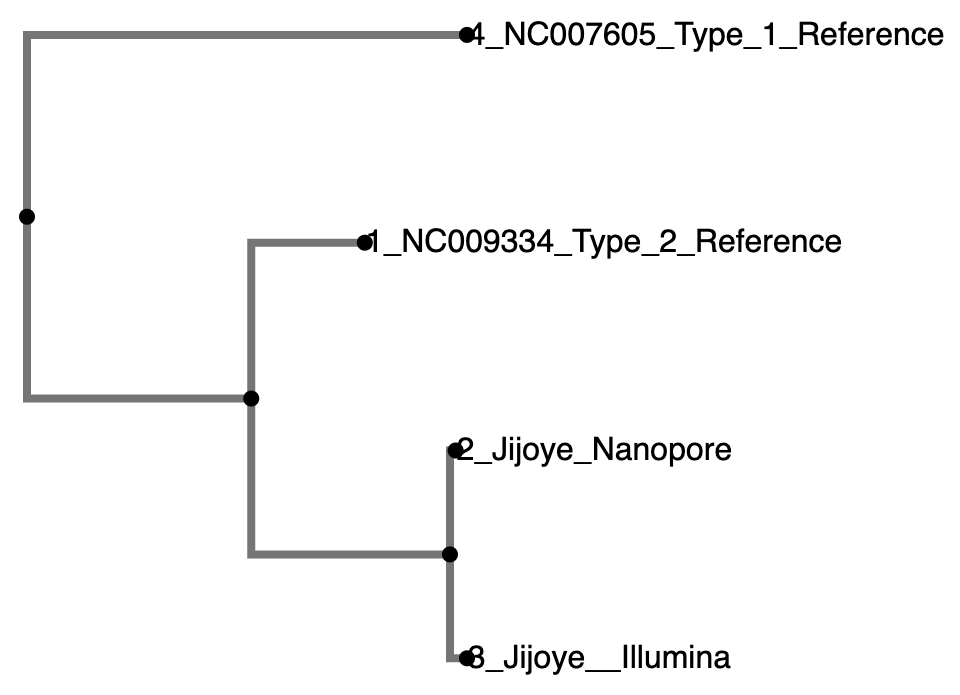

**Supplementary Figure 2. BL719 Phylogenetic Tree**
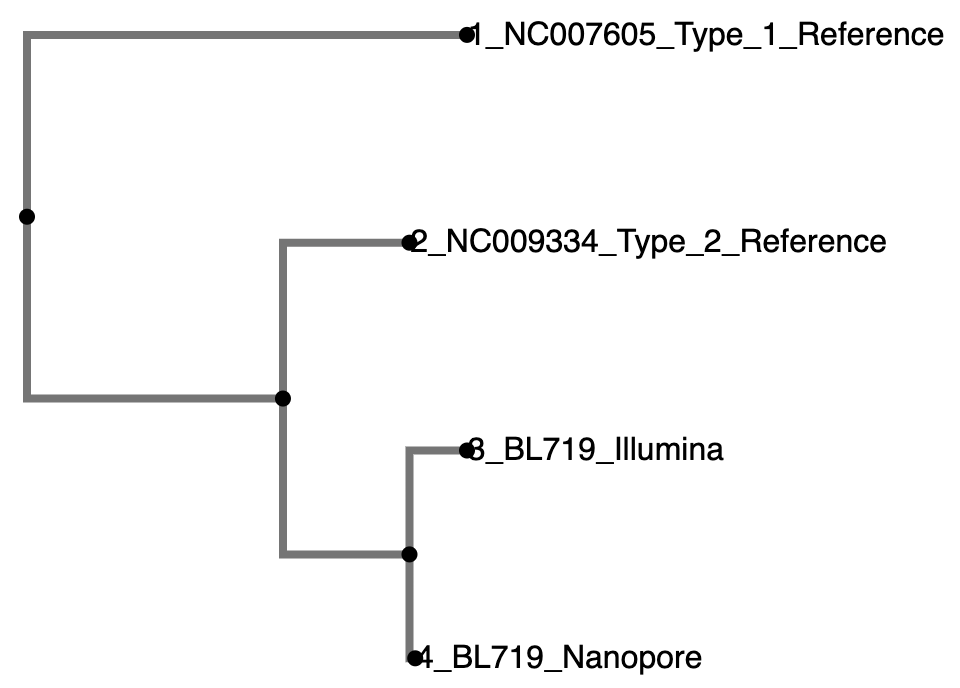

**Supplementary Figure 3. BL725 Phylogenetic Tree**

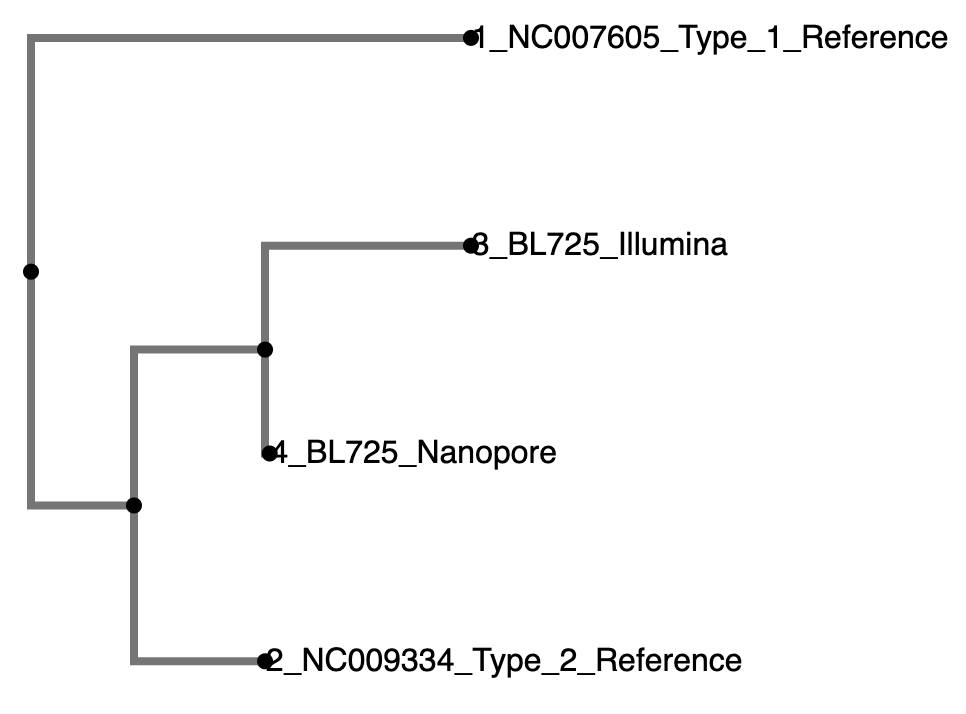

**Supplementary Figure 4. Wewak2 Phylogenetic Tree**

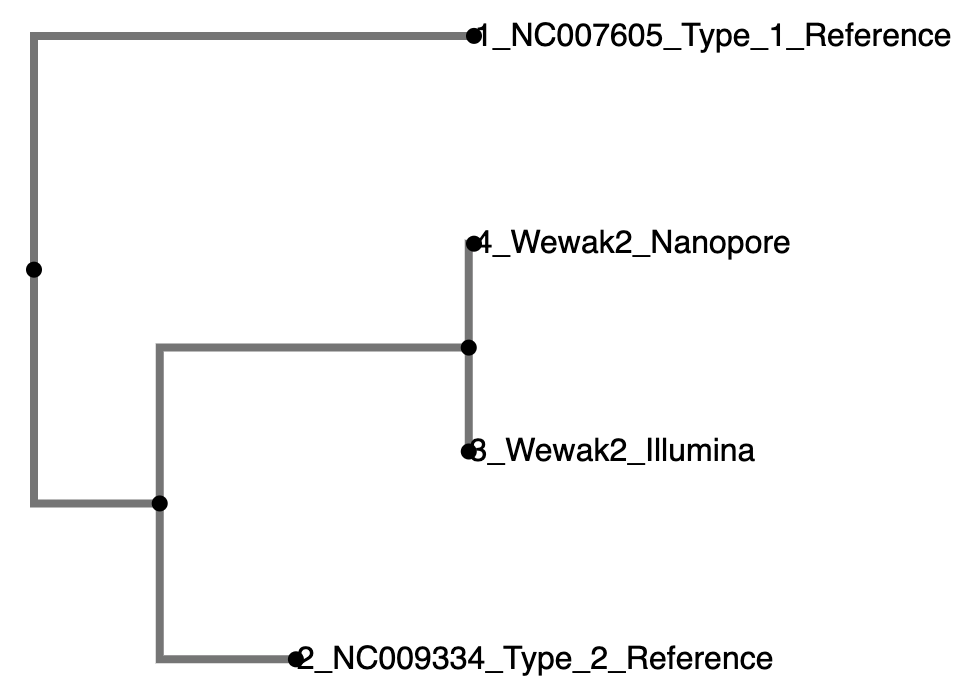

**Supplementary Figure 5. Akata Phylogenetic Tree**

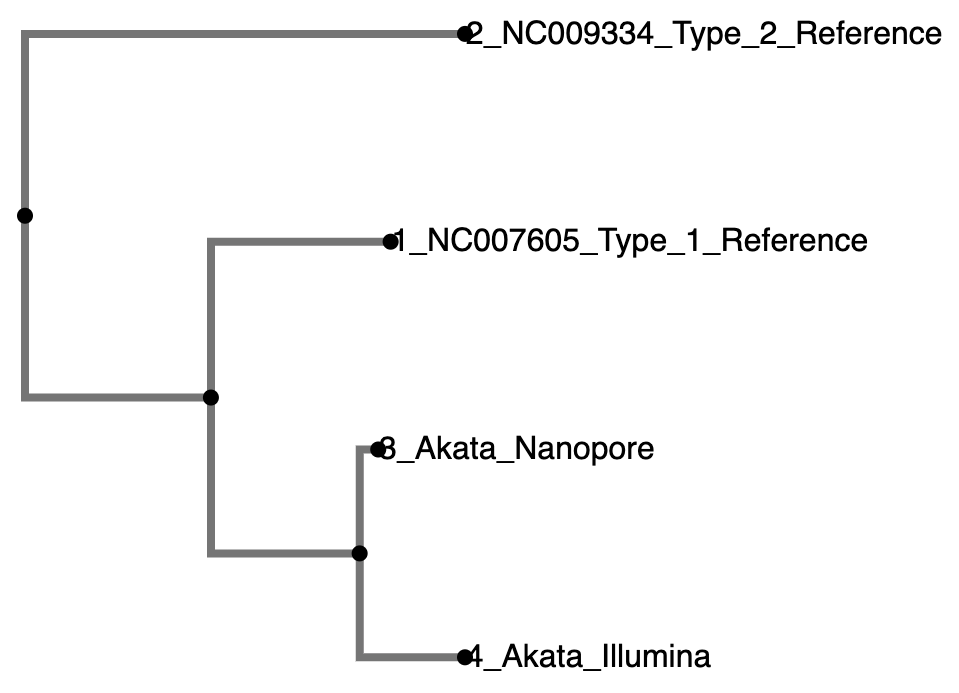

**Supplementary Figure 6. Makau Phylogenetic Tree**

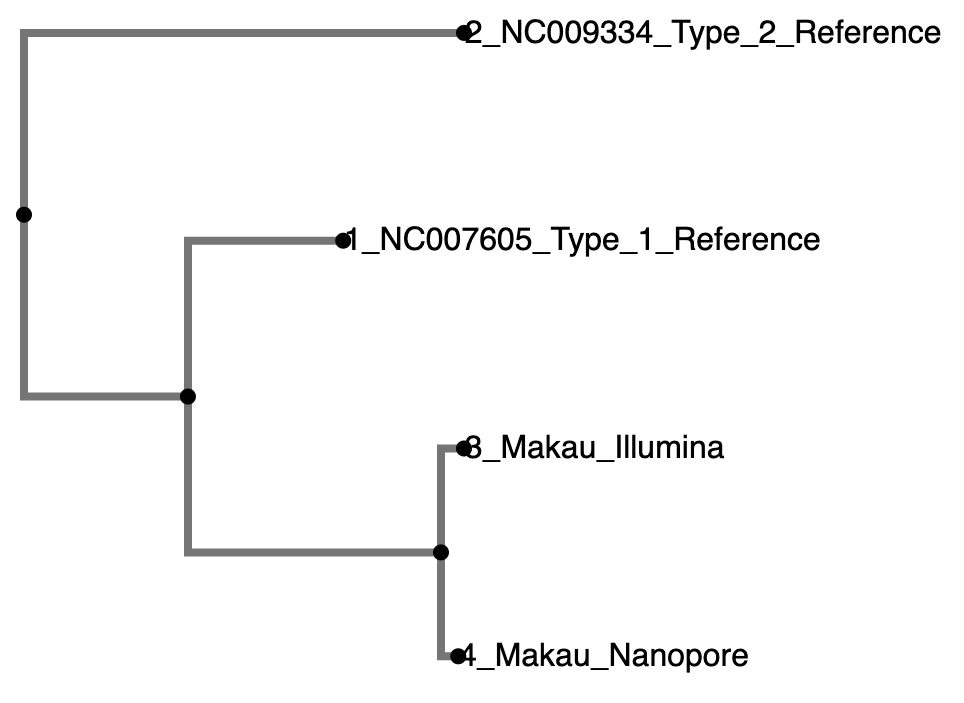

**Supplementary Figure 7. MBL121 Phylogenetic Tree**

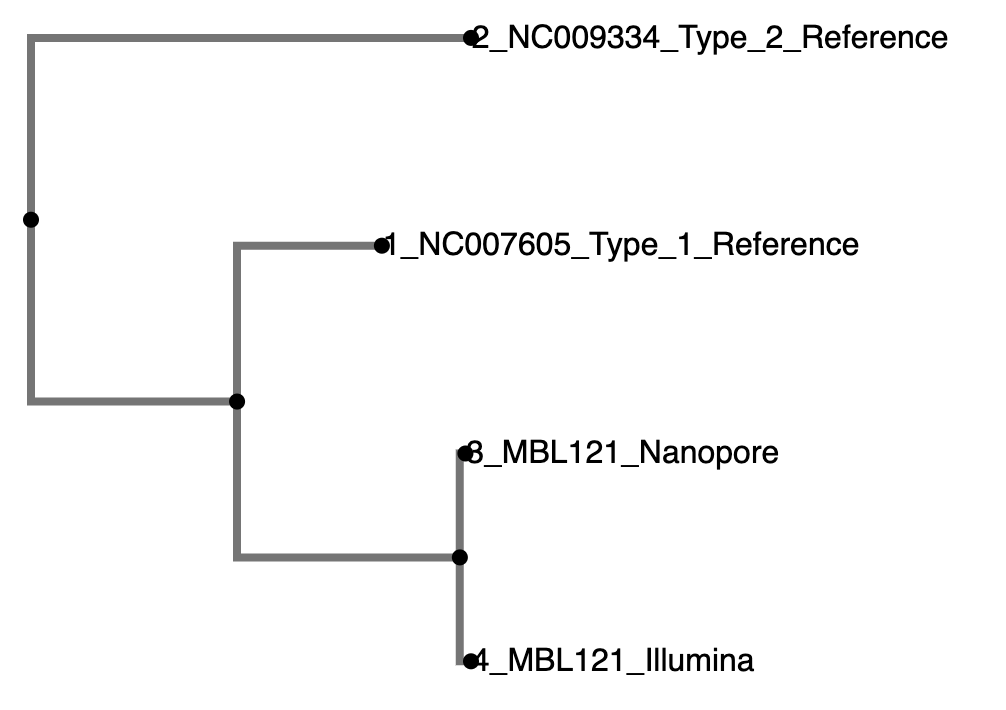

**Supplementary Figure 8. Raji Phylogenetic Tree**

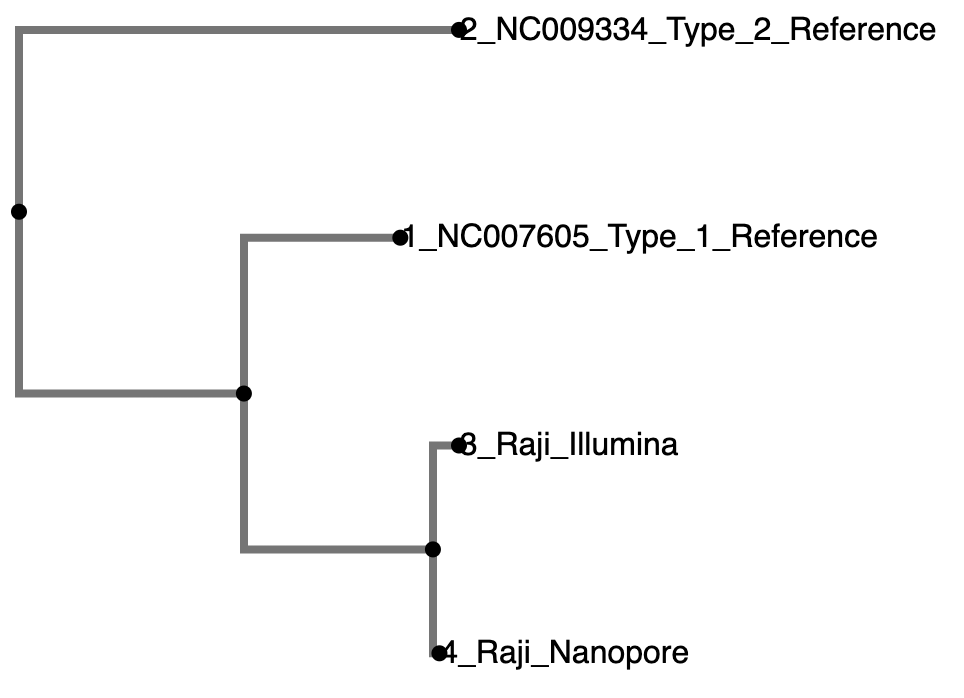

**Supplementary Figure 9. Mutu Phylogenetic Tree**

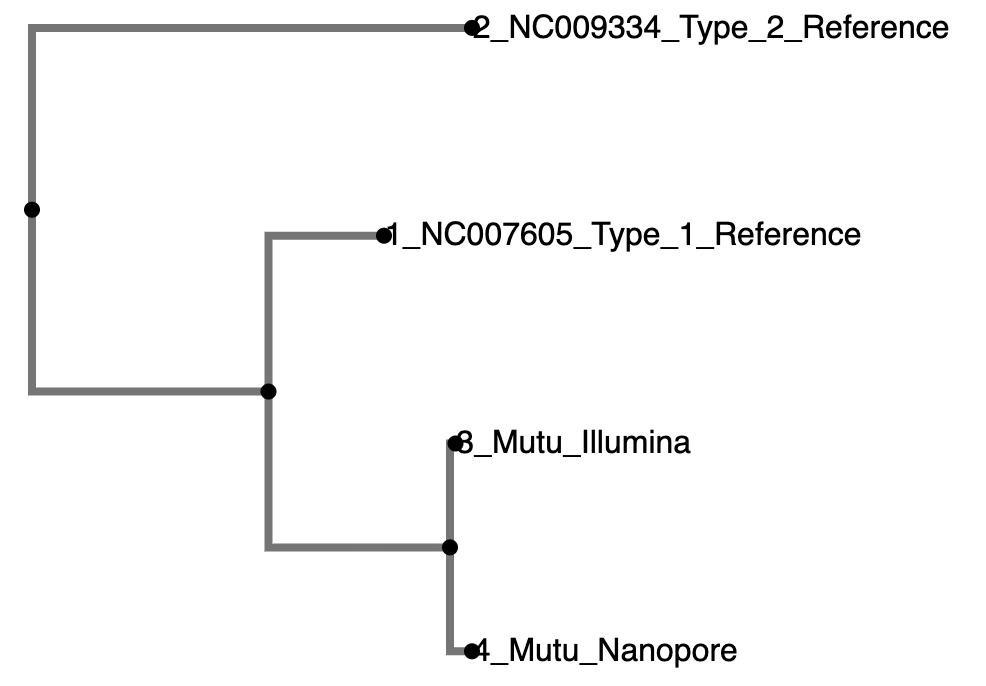

**Supplementary Figure 10. BL720 Phylogenetic Tree**

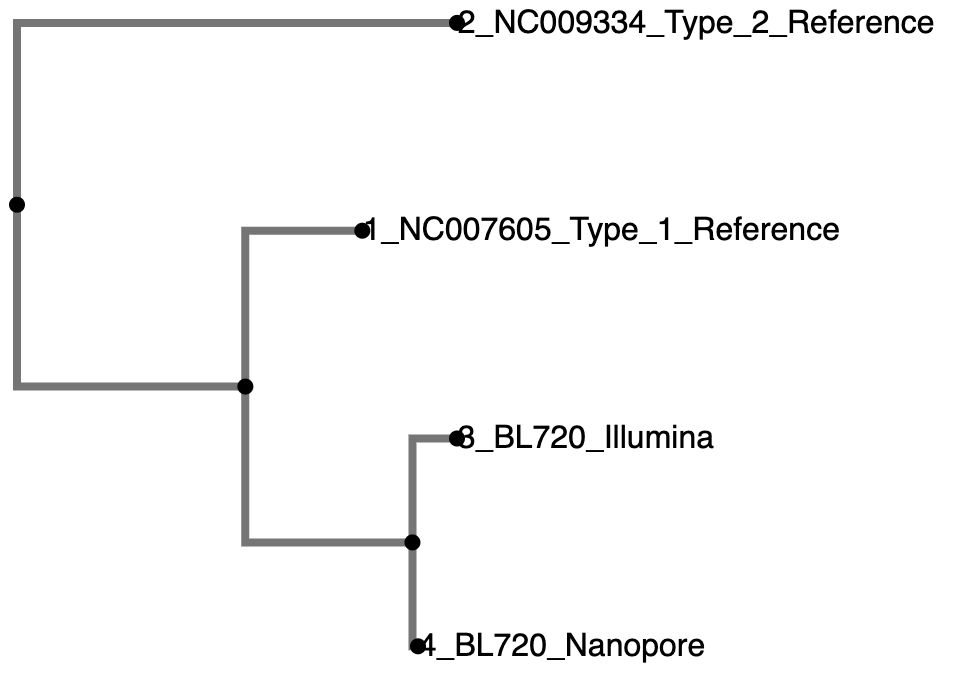

**Supplementary Figure 11. BL740 Phylogenetic Tree**

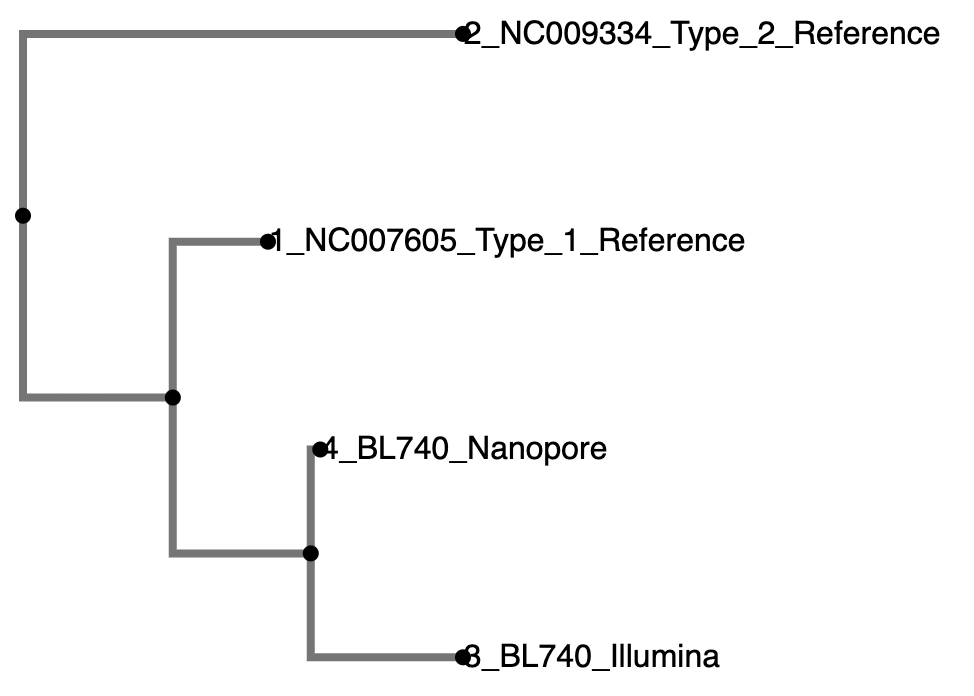

**Supplementary Figure 12. Daudi Phylogenetic Tree**

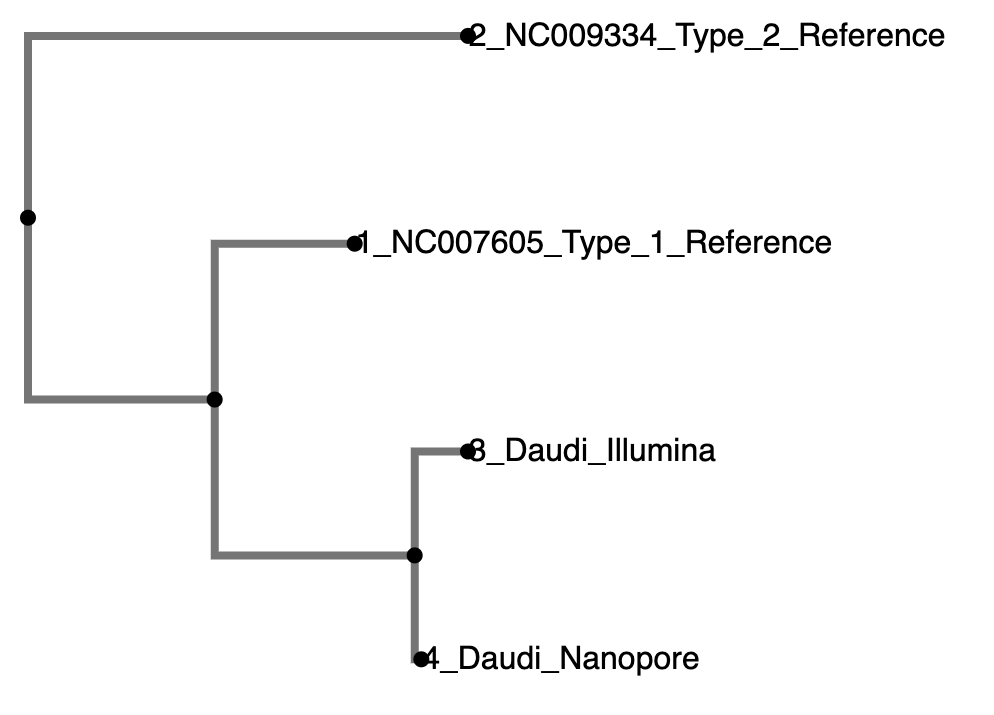

**Supplementary Figure 13. MBL118 Phylogenetic Tree**

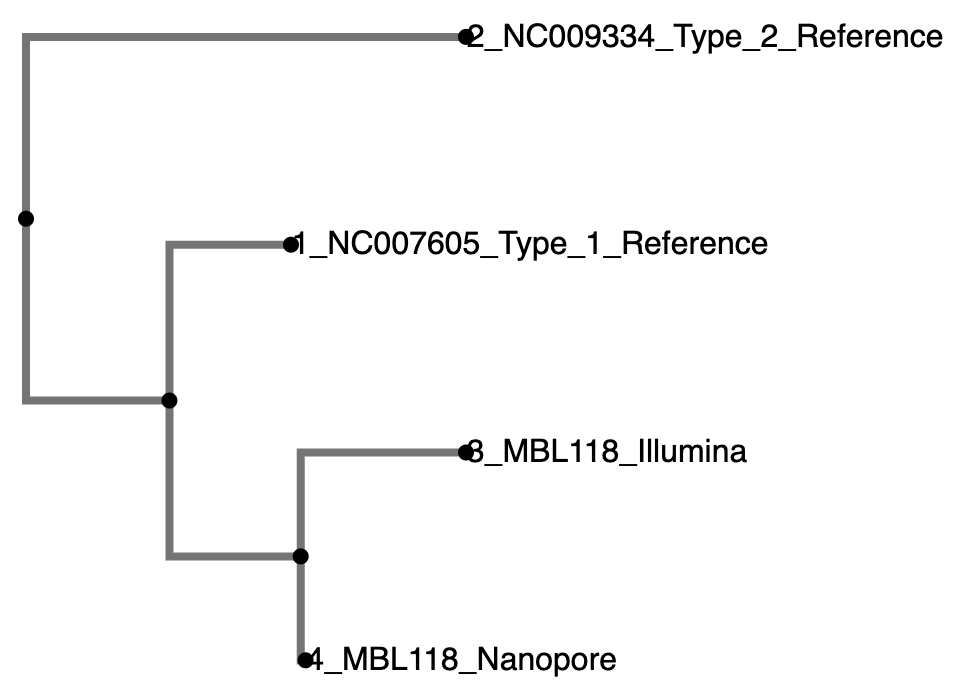

**Supplementary Figure 14. Namalwa Phylogenetic Tree**

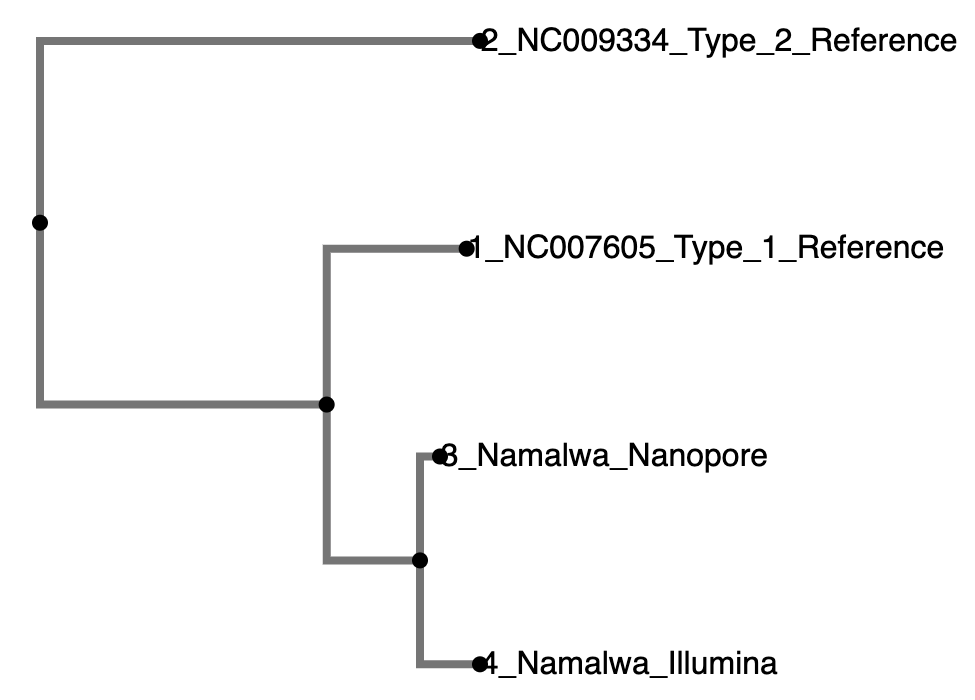
